# Multimodal Neuroimaging Signature of Sleep Problems Predicts Preadolescent Mental Health Trajectories

**DOI:** 10.1101/2025.09.22.25336312

**Authors:** Yulin Wang, Masoud Tahmasian, Sarah Genon, Fateme Samea, Zhihui He, Xinyi Liu, Xu Lei, Simon B. Eickhoff, Debo Dong

## Abstract

Sleep-related problems (SRP) in childhood are common and clinically relevant yet their underlying neural mechanisms and links to future mental health outcomes remain poorly understood. Here, we investigated how distinct dimensions of SRP relate to multimodal brain structure and function in preadolescents, and whether these neural signatures predict trajectories of mental health difficulties. We employed multivariate mapping to investigate the relationship between structural and functional brain network patterns and various dimensions of SRP in the Adolescent Brain Cognitive Development (ABCD) dataset. Moreover, we explored whether and how the identified multimodal brain signatures could predict the trajectory of internalizing and externalizing behavior difficulties over a two-year follow-up. Our multivariate analysis revealed two robust dimensions of SRP: a general sleep disturbance dimension and a hypersomnolence and parasomnia dimension. Each was associated with partially distinct patterns of brain morphology and functional connectivity, consistent with their differential alignment along the hierarchical organization of cortical neurodevelopment maps. However, both dimensions shared common disruptions in the somatosensory, attention, and default mode networks. We further observed that only these neural patterns associated with the general sleep disturbance dimension predict the longitudinal trajectories of internalizing/externalizing symptoms. Our findings enhance the understanding of the neurobiological mechanisms underlying dimensions of SRP in preadolescence and could inform brain-based intervention and treatment programs to improve sleep-related and mental health–related outcomes across development.

## Introduction

Late childhood and early adolescence (i.e., preadolescence) mark the onset of pubertal development, a phase characterized by significant changes in sleep behaviors and the emergence of sleep problems (Friel et al., 2020; Meltzer et al., 2021; Reynolds et al., 2023). During this critical period, sleep problems such as poor sleep quality, nightmares, sleep-talking/walking, abnormal sleep duration, daytime sleepiness, waking after sleep onset, increased sleep onset latency, and difficulty sleeping alone become increasingly prevalent, affecting an estimated 25%–40% of children and adolescents (Cooper et al., 2023; Corkum et al., 2016). In parallel, the rapid rise in screen exposure, including excessive use of smartphones, internet games, and social media, has contributed to digital overload and poses additional risks for sleep disturbances in this age group (Meng et al., 2022). These sleep problems tend to persist into adulthood, often leading to long-term consequences for both physical and mental health (Hysing et al., 2020; Melaku et al., 2019). In particular, sleep disturbances during this period can trigger internalizing symptoms (e.g., anxiety, depressive symptoms) and externalizing symptoms (e.g., attention and behavioral regulation difficulties), while elevating lifelong susceptibility to mental health risks (Scott et al., 2021; Shen et al., 2020; Zhi et al., 2024).

Emerging evidence further highlights the multifaceted role of sleep disturbances in mental health. In addition to their symptomatic impact, sleep disturbances may provide valuable insight into underlying psychopathological mechanisms. Specifically, they not only contribute to the development and maintenance of mental health issues but may also serve as an early marker of vulnerability, aiding in understanding etiology and facilitating early identification and intervention (Abi-Dargham et al., 2023; Cooper et al., 2023; Freeman et al., 2020). For example, a recent study using the Adolescent Brain and Cognitive Development (ABCD) cohort identified sleep disturbances as the strongest predictor of high-risk mental health status among a wide range of behavioral, cognitive, and environmental variables, reinforcing their prognostic significance (Hill et al., 2025). Importantly, large-scale longitudinal analyses further demonstrated that healthy sleep is the most critical lifestyle factor for preventing depression onset, particularly for first episode and treatment-resistant depression in adults (Zhao et al., 2023). However, the neurobiological underpinnings of the association between sleep disturbance and mental health in preadolescence are poorly understood. Thus, addressing this issue and elucidating their associations with the development of internalizing and externalizing symptoms becomes essential.

Disruptions to sleep during preadolescence may have lasting effects on neural circuits underlying attention, emotion regulation, and cognition (Leong & Chee, 2023; Reynolds et al., 2023). Yet, existing studies linking SRP to brain alterations remain fragmented, typically focusing on isolated sleep domains or single neuroimaging modalities (Akbar et al., 2022; Cheng et al., 2021; Isaiah et al., 2021). Although large-scale, multimodal neuroimaging datasets such as ABCD (Hubbard et al., 2020; Volkow et al., 2018) provide the opportunity to capture more complex brain–sleep associations, to date few studies have leveraged them to explicitly integrate structural and functional imaging features in a multivariate framework to address the heterogeneous SRP dimensions. This gap is particularly important given that preadolescent SRP encompass a range of partially overlapping domains—from behavioral difficulties like bedtime resistance to clinically relevant conditions such as parasomnias or sleep breathing problems (McCurry et al., 2024; Meltzer et al., 2021). The high interdependence of these domains suggests the need for dimensional, data-driven approaches that move beyond summary scores or categorical diagnoses (Haritos et al., 2025; Wang et al., 2023). A multimodal, multivariate framework may thus provide a more comprehensive understanding of the neurobiological correlates of SRP and their contribution to emerging psychiatric vulnerability. Unsupervised multivariate techniques, such as partial least squares (PLS), are uniquely suited to advance this integration by identifying latent dimensions that connect interindividual variability in whole-brain structural and functional patterns to behavioral profiles across SRP domains, which can in turn be linked to developmental trajectories or neurotransmitter architectures. These dimensions can then be mapped to psychiatric symptoms, offering novel insights into how specific sleep-related neurobiological alterations contribute to mental health outcomes.

Taken together, this study aimed to disentangle different SRP dimensions with their associated multimodal brain correlates in preadolescence as the primary objective and elucidate their associations with internalizing and externalizing symptoms as a secondary aim. To achieve this, we first employed PLS approach to identify latent dimensions of covariation between structural (cortical surface area, cortical thickness, volume) and functional (resting-state connectivity) brain features and item-level SRP measures derived from the Sleep Disturbance Scale for Children (SDSC) and Child Behavior Checklist (CBCL) in the ABCD dataset (ages 9–11, (Casey et al., 2018)). Analyses followed a split-sample design to ensure replication and generalizability: PLS-derived dimensions were initially modeled in a Discovery subsample (N=3472), internally validated in a Replication subsample (N=1729) (Figure 1). We then contextualized the derived dimensional SRP-related neural correlates within normative neurodevelopment by computing spatial correlations with established cortical developmental maps (Markello et al., 2022). Finally, we evaluated their clinical relevance by testing whether the PLS-derived neural signatures predicted internalizing and externalizing symptoms progression over a two-year period. This integrative strategy provides insight into the neurobiological pathways linking SRP to psychiatric outcomes, highlighting potential neural targets for early intervention for sleep problems, ultimately improving mental health outcomes in this vulnerable population to improve the quality of life and wellbeing in society.

**Fig 1.**
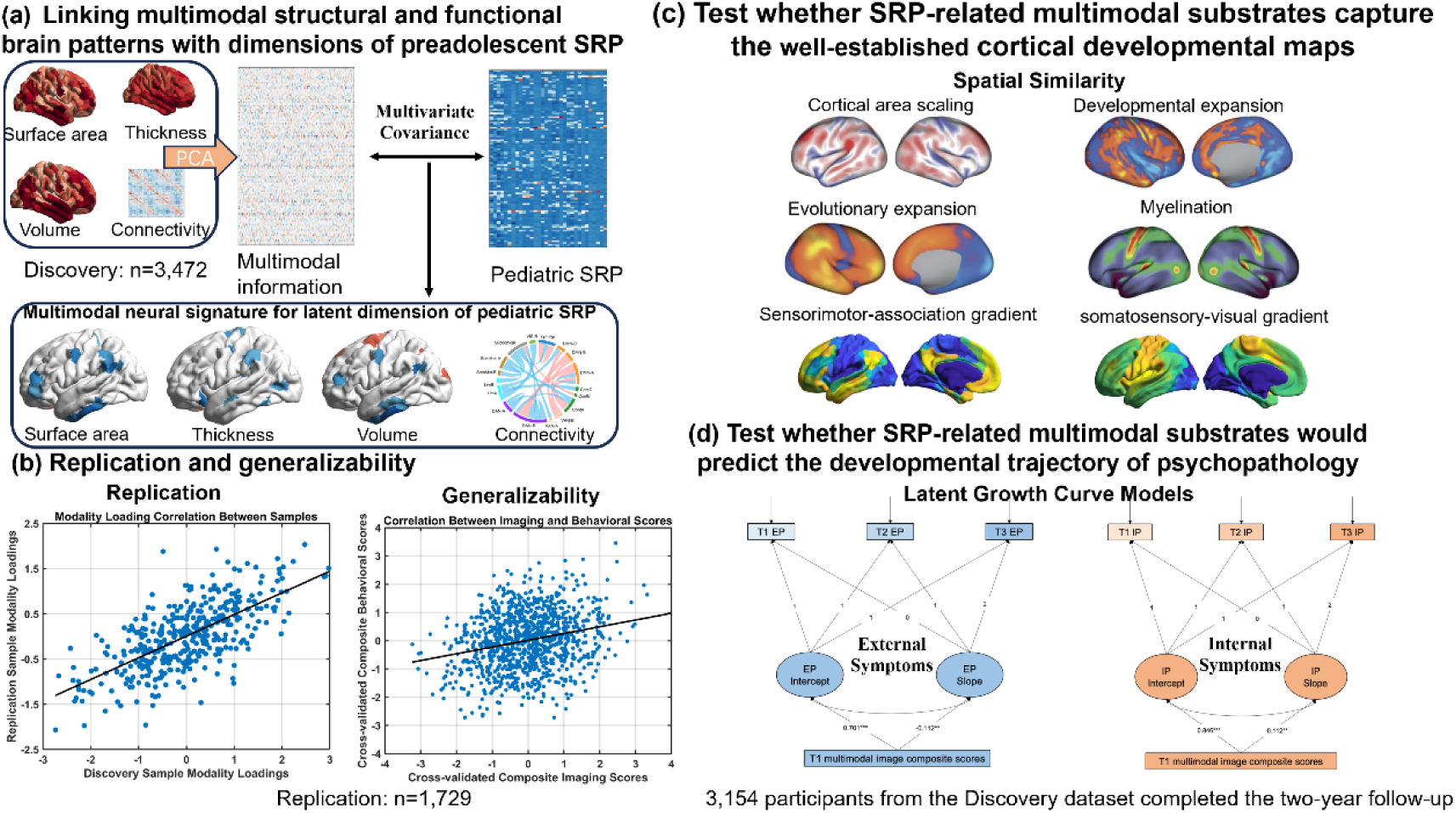
Overview of research questions and main analyses. (a) Linking multimodal structural and functional brain patterns with dimensions of preadolescent SRP in the discovery sample (N=3,472). (b) Replication and generalizability in the replication sample (N=1,729). Replicability was assessed by repeating the PLS analysis in an independent replication dataset and quantifying the correlations of behavioral, RSFC, and structural loadings (surface area, thickness, volume) between the discovery and replication samples. The generalizability of the findings was evaluated by directly applying the model weights from the discovery sample to the replication sample data. (c) Test whether SRP-related multimodal substrates capture the well-established cortical developmental maps. Cortical areal scaling, developmental cortical expansion, evolutionary cortical expansion and cortical myelination maps were adapted from Markello et al. (2022). (d) Test whether SRP-related multimodal substrates would predict the developmental trajectory of psychopathology. T1=Baseline; T2=one year later; T3=Two years later. RSFC, resting-state functional connectivity.

## Results

Our multivariate analysis revealed two distinct and robust SRP-related dimensions: a general sleep disturbances dimension and a hypersomnolence and parasomnia dimension, each covaried with specific structural and functional connectivity signatures as well as shared disruptions mainly involving the somatosensory, attention, and default mode networks. These brain signatures are further contextualized along cortical developmental patterns, as well as functional gradients spanning the somatosensory, attention, and default mode networks in the discovery subsample. Results remained consistent in the replication subsample, indicating generalizability and resilience against variations in analytical parameters. These multimodal signatures mapped onto cortical developmental gradients, suggesting that SRP is embedded within broader neurodevelopmental trajectories. Critically, the identified multimodal neuroimaging signatures associated with the general sleep disturbance dimension can predict the developmental trajectory of internalizing/externalizing behavioral difficulties two years later. Specifically, children with higher multimodal image composite scores (reflecting more pronounced SRP-related neural alterations) tend to experience a slower decrease in their internalizing and externalizing behavioral difficulties over a two-year period.

### PLS model revealed two distinct dimensions, linking SRP and multimodal neuroimaging features

We first identified robust brain-behavior associations using a discovery-replication design. We divided the preprocessed and quality-controlled subsample of the ABCD dataset, which included both structural and resting-state fMRI data, into Discovery (N=3,472) and Replication (N=1,729) subsamples, matched on key demographic and clinical variables (e.g., age, sex, ethnicity, acquisition site, and overall SRP severity). For each participant, we extracted structural and functional neuroimaging features using a 419-node brain parcellation system (400 cortical, 19 subcortical regions) (see **Methods** section for the details). Prior to analysis, both imaging and SRP data were adjusted for relevant covariates, including age, gender, handedness, site, ethnicity, BMI, and socioeconomic status (Dennis et al., 2022; Giddens et al., 2022; Laurent et al., 2020; Morrissey et al., 2020) (Table 1 and Figure S1). Partial Least Squares analysis was then employed to identify patterns of covariance between two high-dimensional matrices: structural and functional imaging features, and 32 SRP items in the ABCD cohort.

**Table 1.** The demographic characteristics in the discovery and replication dataset.

| Variables | Discovery subsample (N = 3472) | Replication subsample (N = 1729) |
| --- | --- | --- |
| Age, mean (SD), years old | 9.94(0.62) | 9.94(0.63) |
| Female (percentage) | 1687(48.59%) | 858(49.62%) |
| Ethnicity |  |  |
| White | 1878 | 953 |
| Black | 452 | 195 |
| Hispanic | 707 | 356 |
| Asian | 77 | 38 |
| Other | 358 | 187 |
| Handedness-Right (percentage) | 2804(80.76%) | 1405(81.26%) |
| BMI, mean (SD) | 18.38(3.78) | 18.86(4.35) |
| Parent education level, mean (SD) | 16.74(2.66) | 16.71(2.68) |

Note. SD, standard deviation; BMI, body mass index.

To reduce data dimensionality before integrating the different imaging modalities, we performed principal component analysis (PCA, 50% explained variation) on each modality (i.e., surface area, cortical thickness, cortical volume, resting state functional connectivity (RSFC)). We retained only the components explaining 50% of the variance within each modality to ensure a balanced contribution across measures and to mitigate the dominant influence of RSFC features. This procedure resulted in a total of 436 components (51 for surface area, 65 for cortical thickness, 58 for cortical volume, and 262 for RSFC), which were then entered into a partial least squares (PLS) analysis along with 32 SRP variables (see Table S1 and Methods for details). Nevertheless, we also present results for different thresholds (see Control analyses).

The PLS analysis identified latent dimensions capturing shared variance between brain features and SRP symptoms. Figure S2 shows the amount of covariance explained by each latent component (LC) in the discovery sample. We focused on components explaining more than 5% of the total covariance; the first three met this criterion (20.09%, 6.99%, and 6.43%) (see **Methods** for the details). After correcting for multiple comparisons, only the first and third components remained statistically significant. To ensure that the identified brain–behavior covariance pattern was not dependent on specific modeling choices, we conducted a series of robustness checks. These included re-running the analysis after adjusting for differences across data collection sites, testing alternative PCA explained variations (30% and 70%) for data reduction, accounting for non-Gaussian distributions in behavioral measures, adding T1w manual quality control scores as an additional control variable, and excluding cortical volume from the imaging inputs. Across all these variations, the overall pattern of association remained highly stable. We assessed the similarity between patterns from each control analysis and the original PLS analysis and found strong consistency across results (r ranged from 0.76 to 0.99, see Table S2 and Methods for details), supporting the robustness of the first and third latent components.

### General sleep disturbances component dimension (LC1)

The first dimension (LC1) indicated a significant brain-behavior association (*r*=0.33, permuted *p* = 2.3 × 10^-3^), linking a distributed neuroimaging pattern with increased severity of preadolescent SRP (Figure 2a). Figure 2b shows the top correlations between LC1’s behavioral loading score and the 20 significant preadolescent SRP measures. Children with higher LC1 behavioral scores tended to experience more frequent and severe sleep difficulties and insomnia symptoms, including trouble falling or staying asleep, waking up tired, and daily sleepiness.

**Figure 2.**
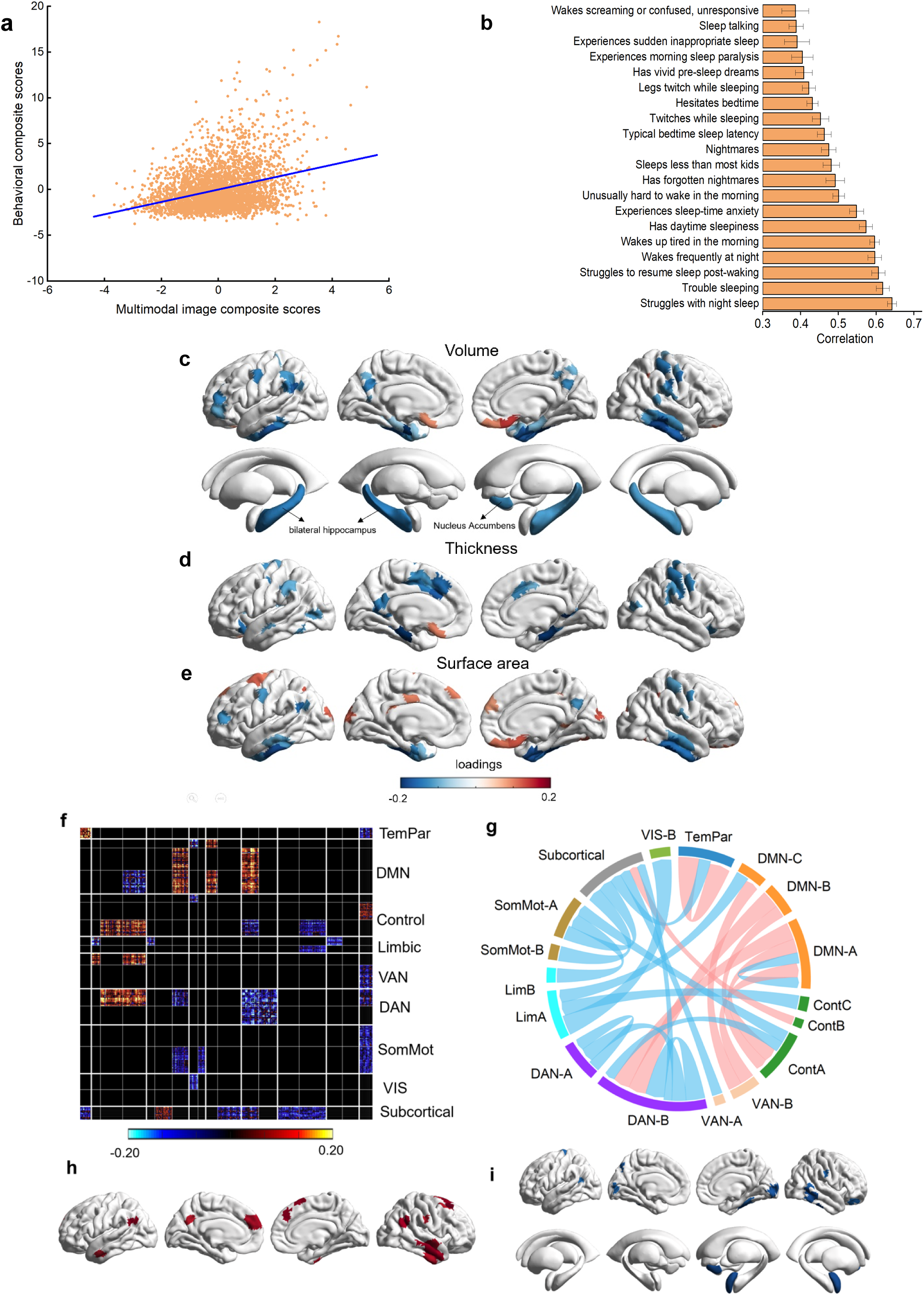
General sleep disturbances component dimension (LC1) a, scatter plots to illustrate the significant association between individual-specific multimodal neuroimaging signatures and behavioral composite scores of participants in LC1 (r =0.33, permuted p = 2.3 x10^-3^). b, significant 20 strongest correlations between participants’ behavioral measures and their behavioral composite scores. Greater loading on LC1 was associated with greater sleep-related problems (SRP). Error bars indicate bootstrapped standard deviation. c, significant volume loadings (after bootstrap resampling and FDR correction q<0.05) associated with LC1. d, significant thickness loadings (after bootstrap resampling and FDR correction q<0.05) associated with LC1. e, significant surface area loadings (after bootstrap resampling and FDR correction q<0.05) associated with LC1. f, significant RSFC loadings (after bootstrap resampling and FDR correction q<0.05). Correlations between participants’ RSFC data and their RSFC composite scores, averaged within and between networks defined by Yeo et al’s 17 network (Yeo et al., 2011) with significant bootstrapped Z scores. g. Chord diagram summarizing significant within- and between-network RSFC loadings. Pink line represents positive correlation while blue line represents negative correlation. h, the top ten nodes with highest degree in the positive network. i, the top ten nodes with highest degree in the negative networks. TempPar, temporoparietal; DMN, default mode network; VAN, ventral attention network; DAN, dorsal attention network; SomMot, somatosensory-motor; VIS, visual network; Cont, control; RSFC, resting-state functional connectivity.

In general, greater SRP (i.e., worse sleep) was associated with volume reductions (Figure 2c) and lower thickness (Figure 2d), while the pattern of surface area associations was more complex, encompassing both increases and decreases (Figure 2e). Specifically, higher SRP was primarily associated with volume reductions mainly in brain regions of the default mode network (DMN) (i.e., middle frontal gyrus, temporal pole, inferior temporal gyrus, bilateral angular gyrus, precuneus cortex, bilateral hippocampus) and the somatosensory network (SMN) (i.e., precentral gyrus and postcentral gyrus). Thickness reductions were mainly observed in the ventral attention network (VAN) (i.e., anterior cingulate cortex, anterior insular cortex, often called the salience network), and SMN (i.e., precentral gyrus and postcentral gyrus) regions. Additionally, surface area reductions were primarily found in the DMN (i.e., precuneus cortex, left angular gyrus, inferior temporal gyrus) and SMN regions (i.e., precentral gyrus and postcentral gyrus), while increases occurred in the superior frontal cortex and ventral medial frontal cortex. Notably, the subgenual anterior cingulate cortex (sgACC) exhibited a common pattern of increase across volume, thickness, and surface area measures.

At the functional connectivity level, severity of SRP was associated with altered large-scale brain network organization (Figure 2f–g). Specifically, we observed increased connectivity between the default mode network (DMN) and several high-order control and attentional systems, including the frontoparietal control network (CON), dorsal attention network (DAN), and ventral attention network (VAN). Within-network connectivity also showed selective increases within the temporoparietal network (TemPar). In contrast, higher SRP was linked to reduced connectivity within the DMN and DAN, between TemPar and subcortical network, between the CON and DAN, decreased connectivity from higher-order systems to limbic/subcortical networks, and decreased connectivity from somatosensory network (SMN) to limbic /subcortical networks.

To better characterize the relative importance of the regions in the obtained significant RSFC pattern, we identified the top ten nodes with highest weighted degree in both the positive and negative networks (Figure 2h–i). Specifically, for the positive network, the top ten nodes with the greatest number of edges were primarily located in the DMN, i.e., bilateral angular gyrus, medial prefrontal cortex, middle temporal gyrus and temporal pole (figure 2h). For the negative network, the top ten nodes with the greatest number of edges were primarily located in the subcortical network, i.e., the amygdala, nucleus accumbens, and SMN (e.g., occipital pole, posterior division of the inferior temporal gyrus, posterior division of the middle temporal gyrus and superior parietal lobule) (figure 2i).

### Hypersomnolence and parasomnia component dimension (LC3)

The third dimension (LC3) distinguished hypersomnolence from parasomnia-related disturbances, indicating a significant association (*r* =0.35, permuted *p* = 2.1 × 10^-4^) between multimodal neuroimaging signatures and behavioral composite scores (figure 3a). Figure 3b shows the top correlations between LC3’s behavioral composite/loading score and the 18 significant pediatric SRP measures. Positive behavioral loadings reflected hypersomnolence symptoms, including excessive daytime sleepiness, prolonged sleep duration, difficulty waking, and pathological sleep inertia (e.g., morning paralysis, Figure 3b). Negative loadings indicated parasomnia symptoms, such as sleep-time anxiety, nightmares, frequent awakenings, and movement-related disturbances (e.g., twitches, sleepwalking; Figure 3b).

**Fig 3.**
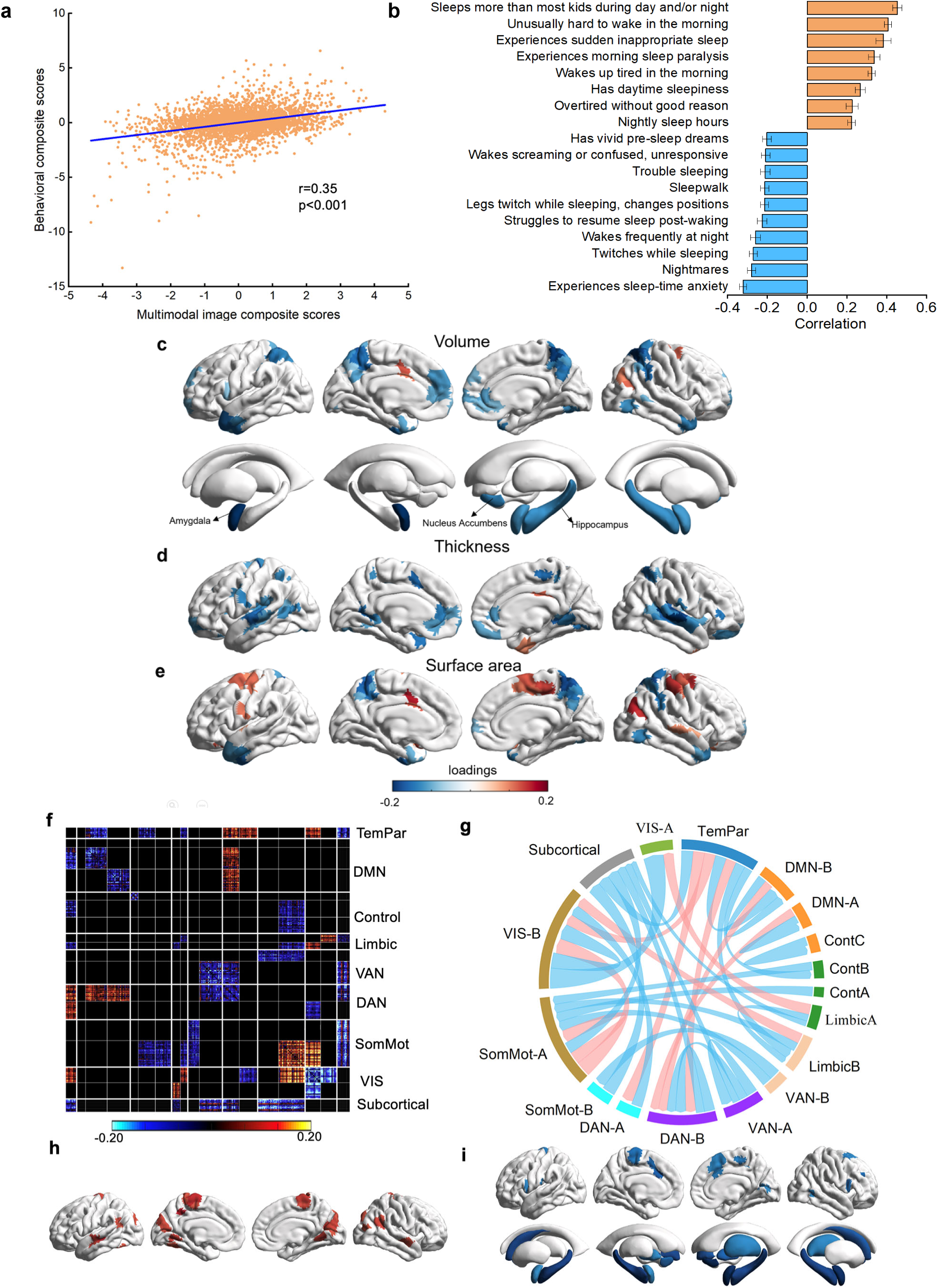
Hypersomnolence-Parasomnia component dimension (LC3) a, scatter plots to illustrate the significant association between individual-specific multimodal neuroimaging signatures and behavioral composite scores of participants in LC3 (r =0.35, permuted p = 2.1 x10^-4^). b, significant 18 strongest correlations between participants’ behavioral measures and their behavioral composite scores. Greater loading on LC3 was associated with greater sleep-related problems (SRP). Error bars indicate bootstrapped standard deviation. Positive behavioral loadings reflected hypo-regulatory symptoms are colored yellow, including sleeps more than most kids, hard to wake in the morning, experience sudden inappropriate sleep, and experience morning paralysis. Negative loadings indicated hyperarousal symptoms are colored in blue, such as sleep-time anxiety, nightmares, frequent awakenings, and movement-related disturbances. c, significant volume loadings (after bootstrap resampling and FDR correction q<0.05) associated with LC3. d, significant thickness loadings (after bootstrap resampling and FDR correction q<0.05) associated with LC3. e, significant surface area loadings (after bootstrap resampling and FDR correction q<0.05) associated with LC3. f, significant RSFC loadings (after bootstrap resampling and FDR correction q<0.05). Correlations between participants’ RSFC data and their RSFC composite scores, averaged within and between networks defined by Yeo et al’s 17 network (Yeo et al., 2011) with significant bootstrapped Z scores. g. Chord diagram summarizing significant within- and between-network RSFC loadings. Pink line represents positive correlation while blue line represents negative correlation. h, the top ten nodes with highest degree in the positive network. i, the top ten nodes with highest degree in the negative networks. TempPar, temporoparietal; DMN, default mode network; VAN, ventral attention network; DAN, dorsal attention network; SomMot, somatosensory-motor; VIS, visual network; Cont, control; RSFC, resting-state functional connectivity.

In general, structural brain differences linked to hypersomnolence and parasomnia component dimension included widespread cortical volume (figure 3c) and thickness reductions (figure 3d), while the pattern of surface area associations was more mixed encompassing increases as well as decreases (figure 3e). Specifically, volume reductions predominantly affected prefrontal and parietal regions (within the control and dorsal attention networks) and subcortical structures (hippocampus, amygdala, nucleus accumbens). Thickness reductions were prominent in attention networks (dorsal/ventral attention networks: superior parietal lobule, anterior cingulate, anterior insula), the somatomotor network (precentral/postcentral gyri), and middle temporal regions of the default mode network. Surface area decreases occurred in the DAN (superior parietal lobule, precuneus) and middle temporal regions, while increases were observed in the SMN (precentral/postcentral gyri).

The third latent dimension, reflecting a hypersomnolence pattern, was associated with distinct alterations in functional network architecture (Figure 3f–g). Children scoring higher on this dimension exhibited (1) reduced functional connectivity within the DMN, attention networks (VAN and DAN), and visual network, and (2) diminished connectivity between subcortical regions and the SMN, DAN and VAN, as well as between the SMN and control/limbic networks, (3) higher functional connectivity within the SMN and between the DMN and DAN. Note that these hypersomnolence brain patterns (e.g., reduced thickness, lower DMN functional connectivity) were inversely related to parasomnia disturbances, which instead correlated with increased cortical thickness and elevated connectivity within the DMN and attention networks.

To better characterize the relative importance of the regions in the obtained significant RSFC pattern of LC3, we identified the top ten nodes with highest weighted degree in both the positive and negative networks (figure 3h–i). Specifically, for the positive network, the top ten nodes with the greatest number of edges were primarily located in the SMN, i.e., precentral gyrus, VN, i.e., lingual gyrus and lateral occipital cortex, and DMN, i.e., middle temporal cortex and angular gyrus (figure 3h). For the negative network, the top ten nodes with the greatest number of edges were primarily located in the SMN, i.e., precentral Gyrus, VAN, i.e., anterior insular and middle cingulate cortex, and subcortical network including hippocampus, amygdala and nucleus accumbens (figure 3i).

### Replicability and generalizability validation

The robustness of the obtained LC1 and LC3 were further ensured with a large replication sample (N=1729) from the ABCD cohort. To do this, we replicated the PLS procedure conducted in the discovery dataset with the replication dataset (see **Methods** for the details). Firstly, the obtained LC1 was largely replicated, evidenced by the high correlation between the behavioral loading scores in the discovery and replication dataset (r= 0.98, p<0.001, figure 4a), between the RSFC loading scores in the discovery and replication dataset (r= 0.18, p<0.0010, figure 4b), between the surface area loading scores in the discovery and replication dataset(r= 0.14, p_spin_=0.042, figure 4c), between the thickness loading scores in the discovery and replication dataset(r= 0.20, p_spin_<0.001, figure 4d), between the volume loading scores in the discovery and replication dataset (r= 0.21, p_spin_<0.001, figure 4e). Secondly, the obtained LC3 was also largely replicated, evidenced by significant correlation between the behavioral loading scores in the discovery and replication dataset (r= 0.80, p∼=0, figure 4f), between the RSFC loading scores in the discovery and replication dataset (r= 0.33, p∼=0, figure 4g), between the surface area loading scores in the discovery and replication dataset (r= 0.34, pspin<0.001, figure 4h), between the volume loading scores in the discovery and replication dataset (r= 0.30, pspin<0.001, figure 4j). However, thickness loadings were not replicated in LC3 (r= 0.08, pspin=0.18, figure 4i).

**Fig 4.**
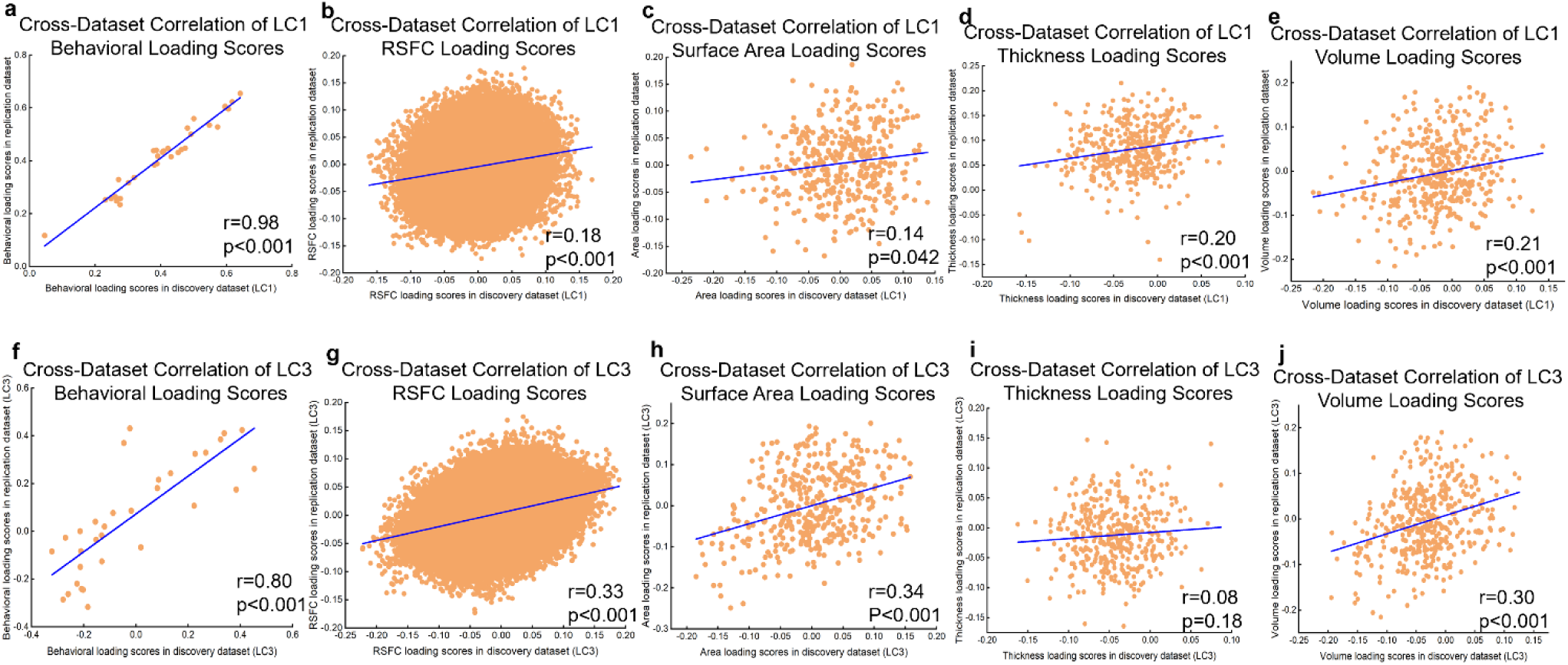
The robustness of the obtained LC1 and LC3 were further ensured with a large replication dataset. a. high correlation between the behavioral loading scores of LC1 in the discovery and replication dataset (r= 0.98, p<0.001). b, the correlation between the RSFC salience scores of LC1 in the discovery and replication dataset (r= 0.18, p<0.001). c, the correlation between the surface area loading scores of LC1 in the discovery and replication dataset (r= 0.14, p_spin_=0.042). d, the correlation between the thickness loading scores of LC1 in the discovery and replication dataset (r= 0.20, p_spin_<0.001). e. the correlation between the volume loading scores of LC1 in the discovery and replication dataset (r= 0.21, p_spin_<0.001). f. high correlation between the behavioral loading scores of LC3 in the discovery and replication dataset (r= 0.80, p<0.001). g, the correlation between the RSFC loading scores of LC3 in the discovery and replication dataset (r= 0.33, p<0.001). h, the correlation between the surface area loading scores of LC3 in the discovery and replication dataset (r= 0.34, p_spin_<0.001). i, the correlation between the thickness loading scores of LC3 in the discovery and replication dataset (r= 0.08, p=0.18). j. the correlation between the volume loading scores of LC3 in the discovery and replication dataset (r= 0.30, p_spin_<0.001). LC, latent component; RSFC, resting-state functional connectivity.

The generalizability of the findings was also evaluated by directly applying the model weights from the discovery sample to the replication sample data. Specifically, the PCA coefficients generated from the discovery cohort’s imaging data were applied to the raw imaging data of the replication cohort. The resulting PCA scores and behavioral data were standardized (z-scored) using the discovery cohort’s mean and standard deviation. Cross-validated composite scores were derived by applying singular value decompositions from the discovery cohort data to the normalized imaging PCA and behavioral data of the replication sample. We then calculated the correlation between the cross-validated composite behavioral and imaging scores to obtain the out-of-sample prediction statistics. Using this approach, we observed significant yet limited out-of-sample prediction accuracy (LC1: r = 0.04; LC3: r = 0.07), with all permuted p-values remaining below 0.001 after 10,000 permutations while accounting for site and multiple comparisons with FDR. These results indicate that the identified brain– behavior dimensions are reproducible across samples, although the predictive strength remains limited, which is similar to prior findings in large-scale, population-based studies of brain– behavior associations.

### Spatial correlation with cortical developmental maps

To further interpret the biological relevance of the identified multimodal brain signatures, we examined whether their spatial topography aligned with established cortical organizational hierarchies (Figure 5a). Such analyses help determine whether SRP-related neural alterations preferentially affect brain regions with specific maturational or evolutionary properties. The following cortical developmental hierarchy maps were investigated based on the literature: 1) cortical area scaling, which reflects how regional surface area scales with total brain size and cognitive capacity, 2) developmental cortical expansion, which captures regional growth trajectories during childhood and adolescence, 3) evolutionary cortical expansion, which highlights regions that have expanded most across primate evolution, 4) cortical myelination, which indicates local myelin content as a proxy for maturational timing, and 5) the recently identified two primary functional gradient maps, derived from the mean RSFC data of the full ABCD sample (N=5,201): represent the sensorimotor-association and somatosensory/motor-visual axis (gradients). These two primary functional gradients also underly the cortical development according to previous studies (Dong et al., 2021). Please refer to the **Supplementary methods** for a detailed derivation of the two primary functional gradients. These cortical developmental maps were then extracted from Neuromap (Markello et al., 2022). First, we obtained the average value of each region of Schafer atlas for each cortical developmental map, i.e., a 400×1 matrix. In the same way, we obtained regional value for unthresholded surface area, thickness, volume, positive and negative RSFC loading maps for each significant component (Figure S3). Finally, we conducted a Spearman correlation analysis between the region loading score and developmental density values calculated for these regions (Dukart et al., 2021). After permutation testing with FDR correction (*q* < 0.05) while adjusting for spatial autocorrelations via spin permutation tests(Alexander-Bloch et al., 2018), significant associations were found for LC1 on the one hand, between the summed positive RSFC network and the cortical area scaling map (figure 5b), the developmental cortical expansion map (figure 5c), the evolutionary cortical expansion map (figure 5d), the cortical myelination map (figure 5e), the sensory-to-transmodal gradient map (figure 5e), and on the other hand, the summed negative RSFC network and the somatosensory/motor-visual gradient map (figure 5i). Also, significant associations were found for LC1 between the thickness loading map and the somatosensory/motor-visual gradient map (figure 5g), and between the volume loading map and the somatosensory/motor-visual gradient map (figure 5h).

**Fig 5.**
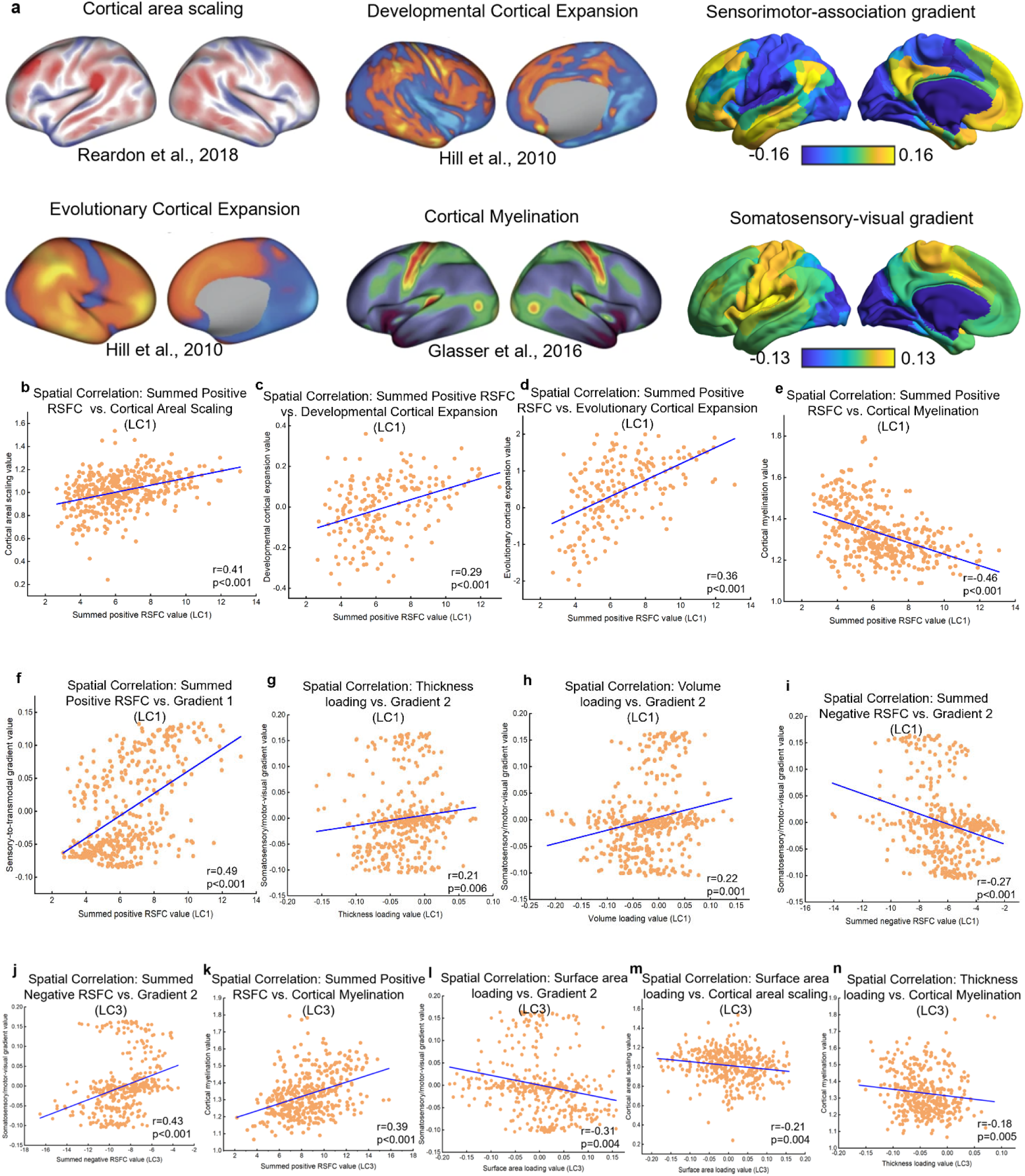
Spatial correlation with cortical developmental maps densities. The multimodal neuroimaging loading scores (weighted) derived from both LC1(b-i) and LC3 (j-n) were spatially correlated with the distribution of cortical developmental maps (a) with Spearman correlational analysis. Specifically, the value of cortical area scaling(b, *r* = 0.41, two sided, *p*<0.001), the value of developmental cortical expansion(c, *r* = 0.29, two sided, *p* <0.001), the value of evolutionary cortical expansion (d, *r* = 0.36, two sided, *p* <0.001), the value of cortical myelination (e, *r* = −0.46, two sided, *p*<0.001), and the value of sensory-to-transmodal gradient (f, *r* = 0.49, two sided, *p*<0.001) were found to be significantly correlated with the summed positive RSFC network derived from LC1; the value of somatosensory/motor-visual gradient (i, *r* = −0.27, two sided, *p*<0.001) were found to be also significantly correlated with the summed negative RSFC network derived from LC1. Also, significant associations were found between the thickness loading value derived from LC1 and the value of somatosensory/motor-visual gradient (g, *r* = 0.21, two-sided, *p* =6.0 x10^-3^), and between the volume loading value derived from LC1 and the value of somatosensory/motor-visual gradient (h, *r* = 0.22, two-sided, *p* =1.0 x10^-3^). The value of the somatosensory/motor-visual gradient was found to be significantly correlated with the summed negative RSFC network derived from LC3(j, *r* = 0.43, two-sided, *p*<0.001); the value of cortical myelination was found to be significantly correlated with the summed positive RSFC network(k, *r* = 0.39, two-sided, *p*<0.001) and the thickness loading(n, *r* = −0.18, two-sided, *p* =5.0 x10^-3^) value derived from LC3. Also, significant associations were found between the surface area loading value derived from LC3 and the value of somatosensory/motor-visual gradient (l, *r* = −0.31, two-sided, *p* =4.0 x10^-3^) as well as the value of cortical area scaling (m, *r* = −0.21, two-sided, *p* =4.0 x10^-3^). LC, latent component. RSFC, resting-state functional connectivity.

For LC3, significant associations were found between the summed negative RSFC network and the somatosensory/motor-visual gradient map (figure 5j), between the summed positive RSFC network and the cortical myelination map (figure 5k), between the surface area loading map and the somatosensory/motor-visual gradient map (figure 5l), as well as the cortical area scaling map (figure 5m), and between the thickness loading map and the cortical myelination map (figure 5n). These results indicate that the significant multimodal neuroimaging patterns derived from both LC1 and LC3 observed in the discovery dataset are spatially correlated with cortical developmental maps.

### Prediction of the developmental trajectory of internalization/externalization problems

We further tested whether the identified multimodal neuroimaging signatures could predict the developmental trajectory of internalizing and externalizing behavioral difficulties. We here applied unconditional Latent Growth Curve Models (LGCM) with Mplus 7.1 to determine the developmental trajectories of the internalizing/externalizing problems, capturing their initial levels (intercept) and changes over time (slope). Then, we incorporated the baseline multimodal neuroimaging composite scores derived from LC1 and LC3 into the conditional LGCM as a time-invariant variable, separately. This approach allowed us to test the predictive role of the identified multimodal neuroimaging composite scores on the developmental trajectories (baseline, 1-year follow-up, 2-year follow-up) of both the internalizing and externalizing problems in preadolescence.

The results for the univariate latent growth curve models are shown in Table S3. Regarding the developmental trajectories of internalizing and externalizing problems, it is indicated that the initial level of internalizing problems was 4.956 (SE=0.094), with a slope of −0.003 (SE=0.041), indicating a general trend of decline over time. However, there was no significant correlation between the initial level of internalizing problems and their subsequent developmental trajectory. In comparison, the initial level of externalizing problems was 4.087 (SE=0.096), with a slope of −0.234 (SE=0.037), showing an overall decreasing trend. Moreover, there was a significant negative correlation (*p*=0.01) between the initial level and the slope of externalizing problems, suggesting that individuals with higher initial levels of externalizing symptoms experienced slower rates of change in externalizing problem behaviors.

Interestingly, multimodal neuroimaging signatures related to general sleep disturbances dimension (LC1) but not LC3 at baseline (T1) significantly predicted both the initial severity and developmental trajectories of internalizing and externalizing problems. Higher composite scores were associated with elevated initial levels of both symptom domains (internalizing: β = 0.846, p < .001, figure 6a; externalizing: β = 0.701, p < .001, figure 6b) and with slower declines over time (internalizing slope: β = –0.112, p = .001, figure 6a; externalizing slope: β = –0.112, p = .001, figure 6b). These findings suggest that preadolescents with more pronounced SRP-related brain alterations may be at increased risk for persistent emotional and behavioral difficulties.

**Fig 6.**
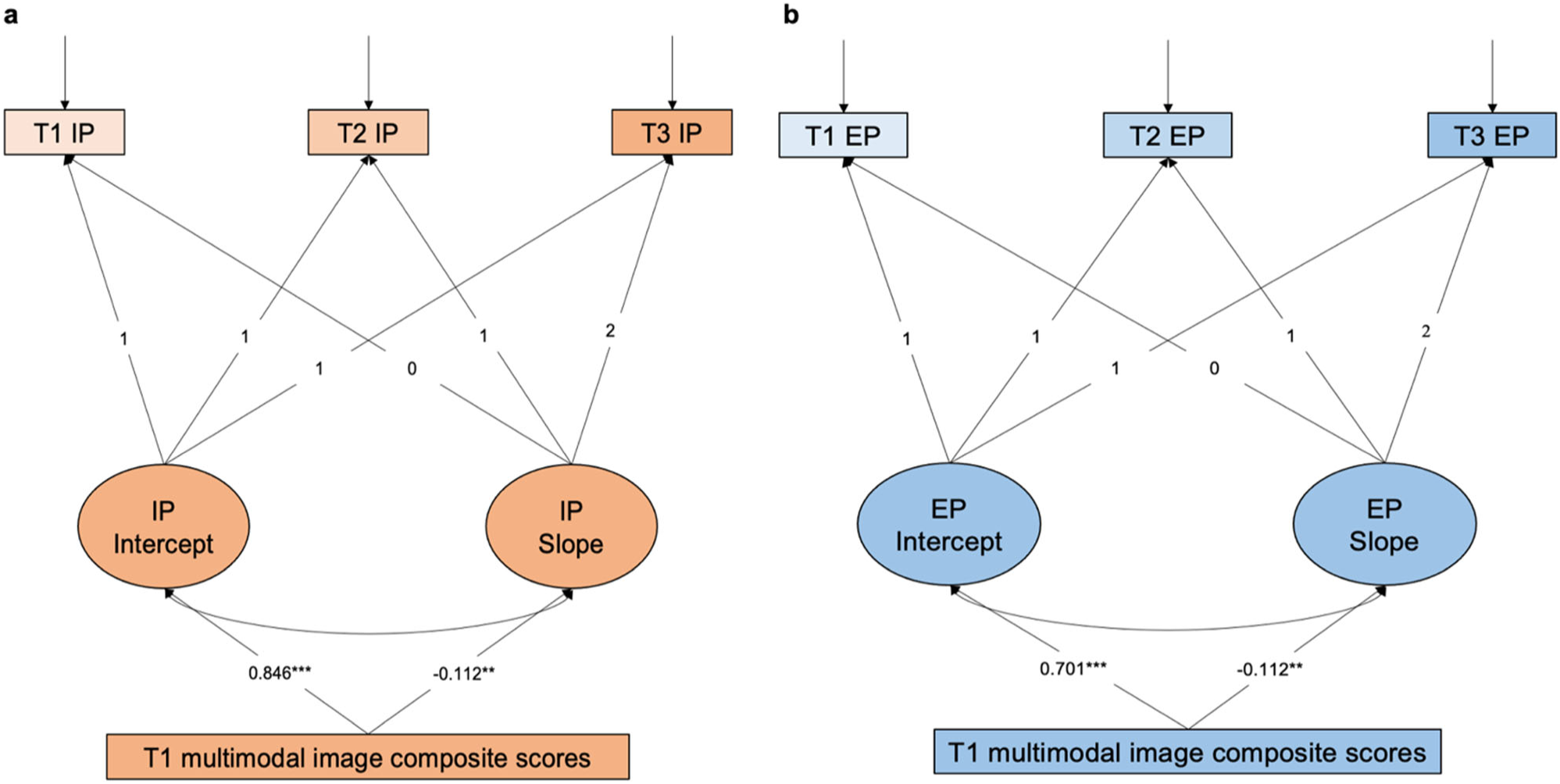
Prediction of the developmental trajectory of internalization/externalization problems. The observed variables at each time point were loaded onto latent intercept and slope factors. To model linear change over time, the time loadings were fixed at 0, 1, and 2, respectively. **a**. general sleep disturbance-related multimodal neuroimaging signatures can predict the developmental trajectory of internalizing problems. The model fit indices demonstrated a good fit to the data (CFI = 1.000, RMSEA = 0.006, SRMR = 0.005, also see table S3). After controlling for age, sex, handedness, sites, ethnicity, parental education, and BMI, the findings revealed that multimodal image composite scores had significant regression coefficients for both the intercept (β = 0.846, p < 0.001;) and the slope (β = −0.112, p = 0.001). **b.** general sleep disturbance -related multimodal neuroimaging signatures can predict the developmental trajectory of externalizing problems. The model fit indices demonstrated a good fit to the data (CFI = 1.000, RMSEA = 0.003, SRMR = 0.005, also see table S3). After controlling for age, sex, handedness, sites, ethnicity, parental education, and BMI, the findings revealed that the general sleep disturbance-related multimodal image composite scores had significant regression coefficients for both the intercept (β = 0.701, p < 0.001) and the slope (β = −0.112, p = 0.001). IP, internalizing problems; EP, externalizing problems; CFI, Comparative Fit Index; RMSEA, Root Mean Square Error of Approximation; SRMR, Standardized Root Mean Square Residual.

## Discussion

The present study employed an integrated multivariate approach, combining structural neuroimaging features with measures of resting-state functional organization to optimize their covariance with various combinations of sleep-related problems in a large community-based preadolescent cohort. We revealed two distinct yet partially overlapping composite SRP dimensions: a general sleep disturbances dimension and a hypersomnolence and parasomnia dimension, both of which covaried with unique morphological and functional connectivity signatures, as well as shared alterations in key brain systems involving the somatosensory, attention, and default mode networks. By contrasting these two latent components, we gain insight into both shared and symptom-specific neural correlates of SRP during late childhood and early adolescence.

The findings associated with the general sleep disturbances dimension, which captures a general dimension of SRP, including difficulties initiating or maintaining sleep, excessive somnolence, and problems with sleep quality, was primarily associated with widespread reductions in brain volume, cortical thickness, and surface area, although an increase in surface area was also observed. These findings are consistent with earlier research implicating sleep-related problems may broadly influence atypical brain morphology during late childhood and early adolescence (Akbar et al., 2022; Cheng et al., 2021; Isaiah et al., 2021). The regions with decreased volume and surface area were primarily located in the DMN and SMN, while the regions with reduced cortical thickness were mainly found in the VAN and SMN. These findings further suggest that sleep disturbances may interfere with the normal maturation of brain regions involved in attentional control and sensory processing, both of which are crucial for cognitive and emotional development during preadolescence (Anastasiades et al., 2022; Dutil et al., 2018; Ricci & Fernandez-Mendoza, 2024). The observed increase in surface area in superior frontal cortex and ventral medial frontal cortex could suggest compensatory mechanisms or the brain’s attempt to adapt to the disruptions caused by sleep-related problems, potentially reflecting neural plasticity (Chen et al., 2022; Porkka-Heiskanen et al., 2013), where the brain adjusts to external and internal factors, but the long-term implications of such changes remain unclear.

On the functional side, the general sleep disturbances dimension was characterized by increased RSFC between the DMN and high-order cognitive networks such as the CON, DAN, and VAN. This indicates a reduced degree of dedifferentiation between the DMN and these cognitive control networks, which impairs the brain’s ability to appropriately switch between internally focused (DMN) and externally directed (CON, DAN, VAN) cognitive processes (Dong et al., 2021; Luo et al., 2024). This reduced segregation may underlie deficits in cognitive flexibility, attention, and emotional regulation, all of which are critical for healthy development during preadolescence (Menon, 2023; Yeshurun et al., 2021). Conversely, LC1 was also characterized by reduced RSFC within the DMN and DAN, weakened connectivity between the CON and DAN, and diminished connections from both higher-order and somatosensory systems to limbic and subcortical regions. These disruptions suggest that SRP impairs not only higher-order cognitive functions but also the integration of basic sensory processing with emotional and motivational systems (Leong & Chee, 2023). Given sleep’s essential role in consolidating sensory information and emotional experiences (Simon et al., 2022), such dysconnectivity may contribute to heightened emotional reactivity and difficulties in processing sensory stimuli, particularly during preadolescence (Chiang et al., 2022). Moreover, reduced intra-network connectivity within the DMN, along with decreased CON–DAN communication, likely reflects impaired coordination across executive and attentional control networks (Menon, 2023; Ribeiro et al., 2024). Together, these alterations may underlie the cognitive and emotional difficulties commonly reported in youths with sleep problems, including concentration deficits, emotional dysregulation, and impaired decision-making (Chiang et al., 2022; Mason et al., 2021; Palmer et al., 2023).

In contrast, the hypersomnolence and parasomnia dimension captured a bidirectional profile of sleep disturbances, distinguishing hypersomnolence (e.g., excessive daytime sleepiness, difficulty waking) from parasomnia symptoms (e.g., nightmares, sleep-time anxiety, movement disturbances). The hypersomnolence subtype may reflect central nervous system under-arousal or delayed circadian phase, commonly observed in preadolescents (Crowley et al., 2018; Lo et al., 2016). The parasomnia subtype aligns with cognitive and physiological hyperarousal models of insomnia, where elevated arousal disrupts sleep onset and maintenance and is often linked to emotional reactivity, nighttime fears, and internalizing symptoms such as anxiety and depression (Gregory & Sadeh, 2016; Riemann et al., 2010). Structurally, the hypersomnolence and parasomnia dimension was associated with volume reductions in prefrontal and subcortical regions (hippocampus, amygdala, nucleus accumbens), along with decreased cortical thickness and surface area in DMN and DAN areas, regions essential for internally focused thinking, attentional control, and emotion regulation. These patterns echo previous findings linking poor sleep with atypical fronto-limbic maturation (Dutil et al., 2018; Telzer et al., 2013). Interestingly, the SMN exhibited a dissociation, showing reduced thickness but increased surface area, possibly reflecting delayed or imbalanced synaptic pruning in circuits involved in bodily awareness and sensory processing (Turan et al., 2025). Functionally, hypersomnolence was characterized by reduced RSFC within DMN, DAN, VAN, and visual networks, as well as weakened connectivity between cortical systems and subcortical regions, consistent with a collapse of prefrontal cortical control, thus impaired emotion regulation (Morales-Muñoz & Gregory, 2023). In contrast, parasomnia showed increased RSFC within the SMN and between the DMN and DAN, reflecting elevated cortical reactivity and attentional hypervigilance suffer from restless nights (Morales-Muñoz & Gregory, 2023). Together, these multimodal findings suggest that the hypersomnolence and parasomnia dimension reflects divergent neural pathways underlying hypersomnolence and parasomnia, advancing our understanding of how SRP phenotypes are rooted in disruptions across arousal modulation, attentional control, and emotional processing networks.

Common to both the general sleep disturbances and hypersomnolence and parasomnia dimension were decreases in cortical thickness in the VAN and SMN, reduced volume and surface area in the DMN, diminished RSFC within higher-order cognitive networks and between high-order and subcortical networks, as well as increased RSFC between the DMN and DAN. Both dimensions also exhibited spatial correspondence with cortical developmental gradients, suggesting a shared disruption in the hierarchical organization of the brain (Dong et al., 2021; Huntenburg et al., 2018). Together, these results suggest that while general sleep disturbances reflect broad neurodevelopmental vulnerability characterized by structural reductions and functional dedifferentiation, the hypersomnolence and parasomnia dimension reflects subtype-specific multi-modal brain imaging signatures along a hyporegulation-to-hyperarousal spectrum. Importantly, both the general sleep disturbances and hypersomnolence and parasomnia dimension signatures aligned with cortical maps of evolutionary and developmental expansion, as well as maps of cortical myelination, reinforcing the relevance of cortical developmental hierarchies in understanding preadolescent SRP-linked brain alterations (Galván, 2020; Mason et al., 2021). These findings underscore the value of a multimodal, dimensional approach to pediatric SRP and provide a potential framework for targeted early interventions that align with the neurobiological underpinnings of specific symptom clusters.

Critically, the multimodal neuroimaging signatures identified in the general sleep disturbances dimension extend beyond capturing current sleep-related symptoms, they predict the developmental course of emotional and behavioral difficulties. Specifically, children with higher general sleep disturbances dimension-derived multimodal image composite scores showed a slower decline in internalizing and externalizing symptoms over time, suggesting that the neural alterations linked to general sleep disturbance reflect enduring developmental vulnerabilities rather than transient disruptions. This interpretation is supported by recent large-scale longitudinal studies showing that sleep disturbances are among the strongest predictors of future psychiatric risk, surpassing other environmental and familial exposures (Hill et al., 2025), with predictive effects mediated through brain connectivity patterns involving the DMN and subcortical systems (Zhi et al., 2024). Together, these findings situate SRP-related neural signatures within broader neurodevelopmental processes linked to long-term mental health outcomes.

Atypical maturation along the sensory-to-transmodal cortical hierarchy, a key organizing principle of brain development, has been widely implicated in multiple psychiatric disorders (Sydnor et al., 2021). Studies of general psychopathology have converged on a disorder-general dysconnectivity pattern, characterized by impaired integration between lower-order sensory networks and higher-order association cortices (Elliott et al., 2018). This dysconnectivity may reflect aberrant differentiation or delayed maturation of transmodal regions, which are among the last to develop and most plastic during preadolescence. Recent work further demonstrates that connectivity signatures associated with general psychopathology align with the same sensory-to-transmodal gradient observed in our general sleep disturbances dimension findings (Royer, Kebets, Piguet, Chen, Ooi, et al., 2024). These parallels suggest that sleep-related disturbances may share neurodevelopmental pathways with broader psychopathological risk. Together, these convergences suggest that SRP-related signatures may not only capture sleep-specific risk but also index transdiagnostic vulnerability to psychopathology.

The present study leveraged a large, community-based sample and employed multivariate fusion methods to identify robust, replicable multimodal neuroimaging signatures of SRP in preadolescents. These signatures generalized across methodological strategies and predicted the longitudinal trajectory of internalizing and externalizing symptoms over two years. Nonetheless, our work has several limitations. First, the use of PLS captures cross-modal covariation but does not establish causal relationships. Future studies should incorporate longitudinal neuroimaging across multiple time points and apply causal modeling approaches to clarify developmental mechanisms. Second, although objective sleep measures such as actigraphy could enhance validity, their limited availability in the ABCD baseline dataset constrained their use here. Despite these limitations, our findings underscore the importance of integrated, multimodal approaches for understanding SRP-related neurodevelopment and provide a foundation for future work aimed at identifying modifiable targets across development.

In summary, our study identified two distinct SRP dimensions in preadolescents, general sleep disturbances and hypersomnolence–parasomnia, each associated with unique morphological and functional connectivity patterns, as well as shared disruptions in brain systems supporting sensory processing, emotion regulation, and cognitive control. These multimodal signatures mapped onto cortical developmental gradients, suggesting that SRP is embedded within broader neurodevelopmental trajectories. Notably, the identified general sleep disturbance dimension related multimodal neuroimaging signatures predicted persistent internalizing and externalizing symptoms over time, highlighting its potential as an early indicator of developmental risk. Our findings not only enhance the understanding of the neurobiological mechanisms underlying dimensions of SRP in preadolescence but also our comprehension of the underlying causes in the connections between sleep and emotional and behavioral problems in preadolescent, which may inform the development of brain-based interventions and treatment programs aimed at improving sleep and mental health outcomes throughout development.

## Online methods

### Participants

#### Discovery and replication datasets

The dataset used in the present study was selected from the Annual Curated Data Release 2.0.1 from the ABCD consortium (https://abcdstudy.org/index.html). The ABCD dataset contains baseline data from more than 11,000 children aged 9 to 11 years recruited from 21 centers throughout the United States with a diverse range of geographic, socioeconomic, ethnic, and health backgrounds (Casey et al., 2018; Garavan et al., 2018). The 21 centers obtained parents’ full written informed consent and the children’s assent. The research procedures and ethical guidelines were followed in accordance with the Institutional Review Boards (IRB) at the University of California, San Diego as well as at each local site.

After excluding participants with incomplete structural MRI, resting-state functional (Somerville et al., 2018) MRI (rs-fMRI), or behavioral data, MRI preprocessing and quality control, and after excluding sites with less than 20 participants, our main analyses included 5201 unrelated children (2643 female (48.93%), 9.94 ± 0.63 years old, 19 sites). Following the strategy reported in previous studies (Royer, Kebets, Piguet, Chen, Ooi, et al., 2024; Xia et al., 2018), we divided this sample into Discovery (N=3472, i.e., 2/3 of the dataset) and Replication (N=1729, i.e., 1/3 of the dataset) subsamples, using randomized data partitioning with both subsamples being matched on age, sex, ethnicity, acquisition site, and overall SRP (i.e., scores of the first principal component were derived from 32 items, which included measures from the SDSC and sleep-related items from the Child Behavior Checklist (CBCL). Figure 2 shows the distribution of these measures in the two samples. See Table 1 for the detailed demographics of each subsample.

### Behavioral assessment

#### Pediatric Sleep Related Problems in the discovery and replication dataset

Pediatric SRP was characterized with the parent-reported Sleep Disturbance Scale for Children (SDSC) as well as the sleep related items in the Child Behavior Checklist (CBCL) (see Table S1 for a complete list of items). Specifically, the SDSC (abcd_sds01) is a comprehensive 26-item questionnaire that evaluates the typical sleep-related behaviors of the child over the previous 6 months (Bruni et al., 1996). The SDSC includes six individual subscales indexing problems with arousal (e.g., sleepwalking, nightmare), sleep hyperhidrosis, sleep breathing disorders, sleep-wake transitions (e.g., limb movements, bruxism), sleep onset and maintenance, and excessive somnolence (e.g., daytime sleepiness). The sleep-relate items from the CBCL (see the follow-up section for more details) mentioned previously were nightmares (cbcl_q47_p), overtired (cbcl_q54_p), sleeps less than most children (cbcl_q76_p), sleeps more than most children during the day and/or night (cbcl_q77_p), trouble sleeping (cbcl_q100_p), wets the bed (cbcl_q108_p), and talks or walks in sleep (cbcl_q92_p). Notably, we did not include the item “talks or walks in sleep” as it overlaps with two items in the SDSC. According to previous studies, the included six sleep-related items in CBCL reflect two different types of pediatric SRP, namely sleep disturbances and sleep difficulties (Friedman et al., 2009; Thomas et al., 2015).

#### Psychopathology Symptoms assessed in the ABCD cohort

The children’s dimensional psychopathology and adaptive functioning were evaluated by the Parent-reported Child Behavior Checklist Scores (abcd_cbcls01). The CBCL includes 113 items, that assess the child’s presentation of various psychopathological symptoms over the preceding six months (Achenbach & Edelbrock, 1983). The CBCL comprises eight syndrome scales contributing to composite scores for internalizing, externalizing, and total problems. The internalizing domain is a broad measure of emotional issues, encompassing withdrawn/depressed, somatic complaints, and anxious/depressed syndrome scales. The externalizing domain provides an overall measure of behavioral problems, including rule-breaking behavior and aggressive behavior syndrome scales. Among the Discovery subsample 3154 had both one- and two-year follow-up data available.

#### MRI acquisition and processing Images for the ABCD cohort

MRI acquisition and processing Images were acquired across 21 sites in the United States with harmonized imaging protocols for 3 Tesla (T) General Electric, Philips, and Siemens scanners (Casey et al., 2018). The imaging acquisition protocol consisted of a localizer, T1-weighted images, four runs of rs-fMRI and T2-weighted images. Detailed MRI scan parameters and preprocessing steps of the T1-weighted, fMRI, and diffusion MRI data have been previously reported and discussed (Hagler et al., 2019). Briefly, T1-weighted imaging was acquired utilizing a 3D protocol with the following parameters: 1mm uniform resolution, repetition time (TR) of 2500 ms, echo time (TE) of 2.88 ms, inversion time (TI) of 1060 ms, a flip angle of 8 degrees, a 256×256 matrix, a field of view (FOV) of 256×256 mm^2^, and 176 sagittal slices. Resting-state MRI data were captured using a multiband EPI sequence characterized by 2.4mm isotropic voxels, TR of 800 ms, TE of 30 ms, a flip angle of 52 degrees, a slice acceleration factor of 6, a 90×90 matrix, an FOV of 216×216 mm^2, and 60 axial slices, along with rapid integrated distortion correction. The resting-state fMRI data collection lasted for 20 minutes, divided into four sessions, during which participants were asked to keep their eyes open and passively view a crosshair on the screen. These minimally pre-processed images were further processed and shared by a recent study(Royer, Kebets, Piguet, Chen, Rong, et al., 2024). Briefly, T1-weighted images were further processed using FreeSurfer 5.3.0 (Fischl et al., 1999). Further processing of functional images included alignment to the T1 images using boundary-based registration, followed by motion filtering, nuisance regression, global signal regression, censoring and bandpass filtering. Finally, preprocessed time series were projected onto FreeSurfer fsaverage6 surface space and smoothed using a 6 mm full-width half maximum kernel. See Supplementary Methods for more details.

#### Extraction of functional and structural features from the ABCD cohort

Resting-state functional connectivity (RSFC) was computed by calculating the Pearson’s correlation among the average timeseries for 400 cortical regions (Schaefer et al., 2018) and 19 subcortical regions (Fischl et al., 2002), resulting in 87,571 connections for each participant. Additionally, Fisher’s R-to-Z transformation was applied to the correlation coefficients to stabilize the variance and normalize the distribution of the correlation values. When computing functional connectivity (FC), censored frames were not taken into consideration. To refine the RSFC data, age, age², sex, handedness, site, ethnicity and head motion (mean framewise displacement [FD]), and image intensity (mean voxel-wise differentiated signal variance [DVARS]) were regressed out. Surface area, thickness, and volume measures were obtained from the same 400 cortical regions (Schaefer et al., 2018). For each parcel-wise structural measure, age, age², sex, handedness, site, and ethnicity were also regressed out, and cortical thickness and volume were additionally adjusted for total intracranial volume, with surface area additionally adjusted for total surface area. Additionally, considering the impact of BMI and family socioeconomic status on children’s sleep and brain development, as well as the significant amount of missing data on household income (261 missing values), we used parents’ educational level as a proxy for family socioeconomic status. Both educational level of parents and BMI were regressed out as covariates.

To reduce data dimensionality before integrating the different imaging modalities, we performed principal component analysis (PCA) on each feature (i.e., surface area, cortical thickness, cortical volume, RSFC). We selected PCA scores based on the number of components explaining 50% of the variance within each data modality before concatenating them (Figure 1). The chosen 50% threshold aimed to balance the relative contribution of each modality, preventing the potentially dominating impact of RSFC features, given their relatively larger number compared to structural features (see Supplemental Methods). Nevertheless, we also present results for different thresholds (see Control analyses). We derived 51, 65, 58, and 262 principal components for surface area, thickness, volume, and RSFC respectively, resulting in a total of 436 components. In the present analysis, we opted to examine brain structural metrics and resting-state fMRI because they constitute the most collected and extensively analyzed imaging phenotypes, allowing for utilization in other independent datasets. Accordingly, they are available for most participants in the ABCD study. Finally, from a neuroscience standpoint, they offer valuable insights into the configuration of the brain’s intrinsic gray matter networks.

#### Partial least squares analysis

Partial least squares (PLS) correlation analysis (McIntosh & Lobaugh, 2004; McIntosh & Mišić, 2013) was used to identify latent variables (LVs) that optimally relate children’s SRP (Y), as indexed by the 32 sleep related items in the ABCD cohort, to the obtained 436 structural and functional imaging PCA features (Xpca). PLS maximizes the covariance between two data matrices by linearly projecting the behavioral and imaging data into a low-dimensional space. Briefly, after z-scoring *Xpca* and *Y* (across all participants), a cross-covariance matrix was computed between the imaging and behavioral data matrices. Singular value decomposition was then applied to this cross-covariance matrix, resulting in three matrices: a diagonal matrix containing singular values, and two matrices containing imaging and behavioral weights (V and U). Participants’ imaging and behavioral composite scores (Lx and Ly) were computed by multiplying their original imaging and behavior data with their respective weights. The contribution of each variable to the LVs was determined by computing Pearson’s correlations between participants’ composite scores and their original data, referred to as loadings. The covariance explained by each LV was computed as the squared singular value divided by the squared sum of all singular values.

Statistical significance of the LVs was assessed using permutation testing, specifically 10,000 permutations accounting for sites, over the singular values of the first three LVs (only components explaining more than 5% of the variance were considered, the explained variances for the first three LVs were: 20.09%, 6.99%, and 6.43%, respectively). A false discovery rate (FDR) correction with a threshold of q < 0.05 was applied to the permuted p-values of the three LVs to control for multiple comparisons. For the significant LVs, we obtained each modality loading (surface area, thickness, volume and RSFC) by computing Pearson’s correlation between the PCA-combined multimodal composite scores and each modality data. Loading stability was determined using bootstraps, where data were sampled 5,000 times with replacement among participants from the same site. Bootstrapped z-scores were computed by dividing each loading (surface area, thickness, volume, and RSFC) by its bootstrapped standard deviation. To limit the number of multiple comparisons, the bootstrapped *r*sFC were averaged across edge pairs within and between 18 networks before computing z-scores.

To assess the relative contribution of each modality imaging data to the image loadings of each LC, we calculated Pearson’s correlation coefficients between the full imaging composite scores (Lx), which were obtained by multiplying the PCA-imaging data (Xpca) by the image weight (V), and the composite scores specific to each imaging modality. To derive these modality-specific composite scores, we first isolated the contribution of each imaging type by setting all other modality saliences to zero while retaining only the relevant saliences for the modality under consideration. For instance, when evaluating the significance of surface area loadings, we focused on the first 51 rows of the salience vector V (corresponding to the number of surface area features remaining after PCA), zeroing out the remaining rows, referred to modality-specific saliences (V_modality_). Then, we obtained modality-specific composite scores by multiplying the imaging data (Xpca) by the modality-specific saliences (V_modality_). We found the relative contribution of each modality imaging data is 0.56 for surface area, 0.47 for thickness, 0.61 for volume and 0.68 for RSFC to the image loadings of each LC1, 0.61 for surface area, 0.50 for thickness, 0.63 for volume and 0.62 for RSFC to the image loadings of each LC3.

#### Control and Reliability Analyses

We conducted several control analyses to assess the reliability of our findings. In the original analysis with the discovery dataset, we employed linear regression to account for site effects. As a control analysis, we also used ComBat algorithm (Fortin et al., 2018)to mitigate the impact of site variability Additionally, we performed PLS analyses while retaining principal components that accounted for 30% and 70% of the variance within each imaging modality. As overall SRP behavioral measures included in the PLS analysis were non-Gaussian distribution (as shown in Figure 2), to exclude the potential effect on the robustness of LVs, we used quantile normalization to improve the Gaussian distributions of the behavioral data and re-computed PLS between the normalized behavioral and multimodal neuroimaging data. We further included T1w manual quality control scores as a covariate within a multivariable linear regression model, designed to adjust for possible interferences present in the structural imaging data. Given that cortical volume is determined by the combination of thickness and surface area, we conducted an additional PLS analysis, excluding cortical volume as an imaging metric. To assess the robustness, we computed Pearson’s correlations between each MRI modality (or behavioral) loading obtained in each control analysis and each MRI modality (or behavioral) loading from the original PLS analysis.

#### Replicability and generalizability validation

To internally validate the obtained latent variables (LVs) or dimensions from the discovery dataset, we replicated the partial least squares (PLS) analysis described in the “Partial Least Squares Analysis” sections using the replication dataset (N=1729, comprising one-third of our full sample). We then calculated Pearson’s correlation coefficients between the behavioral loading scores, RSFC loading scores, surface area loading scores, thickness loading scores, and volume loading scores from the discovery and replication datasets, respectively.

The generalizability of the findings was also evaluated by directly applying the model weights from the discovery sample to the replication sample data. Specifically, the PCA coefficients generated from the discovery cohort’s imaging data were applied to the raw imaging data of the replication cohort. The resulting PCA scores and behavioral data were standardized (z-scored) using the discovery cohort’s mean and standard deviation. Cross-validated composite scores were derived by applying singular value decompositions from the discovery cohort data to the normalized imaging PCA and behavioral data of the replication sample. We then calculated the correlation between the cross-validated composite behavioral and imaging scores to obtain the out-of-sample prediction statistics. Statistical significance was assessed using 10,000 permutations, controlling for site effects and correcting for multiple comparisons using the false discovery rate (FDR).

#### Spatial Correlation with cortical developmental maps

We further test whether the multimodal neuroimaging loading scores were spatially correlated with the distribution of cortical developmental maps. Based on the literature, the following cortical developmental maps: 1) cortical areal scaling(Reardon et al., 2018), 2) developmental cortical expansion(Hill et al., 2010), 3) evolutionary cortical expansion(Hill et al., 2010), 4) cortical myelination (Glasser et al., 2016) These cortical developmental maps were then extracted from Neuromap (Markello et al., 2022). Additionally, we examined whether the multimodal neuroimaging loading scores were spatially correlated with two recently identified primary functional gradients (i.e., sensory-to-transmodal and somatosensory/motor-visual gradient) derived from the RSFC data of our full ABCD sample. These two primary functional gradients also underly the cortical development according to previous studies (H. Dong et al., 2021). Please refer to the **supplementary methods** for a detailed derivation of the two primary functional gradients. Then, we obtained the average value of each region of Schafer atlas for each cortical developmental density map, i.e., a 400×1 matrix. In the same way, we obtained regional value for unthresholded surface area, thickness, volume, positive and negative RSFC loading maps (Figure S3). Finally, we conducted a Spearman correlation analysis between the region importance score and developmental density values calculated for these regions(Dukart et al., 2021). To establish the statistical significance of a spatial correlation against chance, we conducted spatial permutation tests to obtain a null distribution of correlation coefficients for region importance scores and developmental density values extracted from 10, 000 permutations while accounting for the spatial autocorrelation of brain regions(Burt et al., 2020; Markello & Misic, 2021). The p-value was determined by empirically observed spatial similarity values compared to the null distribution. Significance level was set at FDR corrected p<0.05.

#### Prediction of the developmental trajectory of internalization/externalization problems

We further tested whether the identified SRP-related multimodal neuroimaging signatures could predict the developmental trajectory of internalizing and externalizing behavioral difficulties. Using Mplus 7.1, we applied unconditional Latent Growth Curve Models (LGCM) (Duncan et al., 2013; Willett & Sayer, 1994) to determine the developmental trajectories of the internalizing/externalizing problems, capturing their initial levels (intercept) and changes over time (slope). We then incorporated the baseline multimodal neuroimaging composite scores derived from our PLS model into the conditional LGCM as a time-invariant variable. This approach allowed us to test the predictive role of these multimodal neuroimaging composite scores on the developmental trajectories (baseline, 1-year follow-up, 2-year follow-up) of internalizing and externalizing problems.

## Data Availability

The ABCD data are publicly accessible through the NIMH Data Archive (NDA) (https://nda.nih.gov/abcd).

## Code Availability

The image preprocessing pipeline is documented at https://github.com/ThomasYeoLab/CBIG/tree/master/stable_projects/preprocessing/CBIG_f MRI_Preproc2016. Study-specific preprocessing code can be found here: https://github.com/ThomasYeoLab/ABCD_scripts. The code for main analyses can be found here: https://github.com/valkebets/multimodal_psychopathology_components. Additionally, the code for PLS analyses is available at https://github.com/danizoeller/myPLS (Zöller et al., 2017)(Zöller et al., 2017). The code for spatial permutation testing can be found at https://github.com/frantisekvasa/rotate_parcellation (Váša et al., 2018)(Váša et al., 2018). Code for ComBat algorithm is available at (https://github.com/Jfortin1/ComBatHarmonization (Fortin et al., 2018).

## Funding

This work was supported by the National Natural Science Foundation of China (Grant No’s. 82202247 and 32300861) and Natural Science Foundation of Chongqing Municipality (No. 2023NSCQ-MSX4446).

## Acknowledgements

We thank Dr. Valeria Kebets from the Montreal Neurological Institute, McGill University, for sharing the processed ABCD multimodal neuroimaging features. Data used in the preparation of this article were obtained from the Adolescent Brain Cognitive DevelopmentSM (ABCD) Study (https://abcdstudy.org), held in the NIMH Data Archive (NDA). This is a multisite, longitudinal study designed to recruit more than 10,000 children age 9-10 and follow them over 10 years into early adulthood. The ABCD Study® is supported by the National Institutes of Health and additional federal partners under award numbers U01DA041048, U01DA050989, U01DA051016,<colcnt=10> U01DA041022, U01DA051018, U01DA051037, U01DA050987, U01DA041174, U01DA041106, U01DA041117, U01DA041028, U01DA041134, U01DA050988, U01DA051039, U01DA041156, U01DA041025, U01DA041120, U01DA051038, U01DA041148, U01DA041093, U01DA041089, U24DA041123, U24DA041147. A full list of supporters is available at https://abcdstudy.org/federal-partners.html. A listing of participating sites and a complete listing of the study investigators can be found at https://abcdstudy.org/consortium_members/. ABCD consortium investigators designed and implemented the study and/or provided data but did not necessarily participate in the analysis or writing of this report. This manuscript reflects the views of the authors and may not reflect the opinions or views of the NIH or ABCD consortium investigators. The ABCD data repository grows and changes over time. The ABCD data used in this report came from http://dx.doi.org/10.15154/1504041.

## Supplementary Materials

### Supplemental Methods

#### MRI processing for ABCD dataset

##### a) Structural MRI Analysis

The T1-weighted images were first corrected for gradient warp and bias field distortions. They were then resampled to a canonical brain space with isotropic voxel dimensions, aligning to a standard reference brain (Hagler et al., 2019). Further processing was conducted using FreeSurfer version 5.3.0, which included the generation of cortical surface meshes for each subject and their alignment to a unified spherical coordinate system (Fischl, Sereno, & Dale, 1999; Fischl, Sereno, Tootell, et al., 1999). Participants whose data failed the recon-all quality checks were not included in the study.

##### b) Functional MRI (fMRI) Analysis

The ABCD initial phase of processing encompassed motion correction, B0 distortion correction, and the rectification of gradient non-linearities, culminating in the resampling of data to achieve isotropic resolution. The subsequent steps for fMRI data included: (1) the elimination of initial acquisition frames (Hagler et al., 2019); (2) the co-registration of structural and functional datasets using boundary-based registration (Greve & Fischl, 2009), with exclusion of runs exhibiting a boundary-based registration cost exceeding 0.6. Measures of framewise displacement (FD, (Jenkinson et al., 2002)) and voxel-wise differentiated signal variance (DVARS, (Power et al., 2012)) were derived using fsl_motion_outliers, with frames exceeding FD of 0.3 mm or DVARS of 50, along with one frame before and two frames after, flagged as outliers and subsequently censored. Data segments with fewer than five consecutive frames after censoring were also removed (Kong et al., 2019). Runs with a censoring rate above 50% and/or maximum FD exceeding 5 mm were excluded. Participants with less than 4 minutes of valid data were also excluded. Nuisance variables, comprising the global signal, six motion parameters, and the average signals from ventricular and white matter regions along with their time derivatives, were regressed from the fMRI time series, with censored frames excluded from this regression. Data were then interpolated across censored frames via least squares spectral estimation (Power et al., 2014). A bandpass filter, with a range of 0.009 Hz to 0.08 Hz, was applied to the data. The final step involved projecting the preprocessed time series onto the FreeSurfer fsaverage6 surface space and applying a 6 mm full-width half maximum smoothing kernel.

#### The derivation of the two primary functional gradients

In order to examine whether the pediatric SRP related multi-model neuroimaging loading scores were spatially correlated with the two primary functional gradient of brain organization, we used diffusion map embedding (Coifman et al., 2005), a nonlinear dimensionality reduction method, on RSFC data. This technique places strongly interconnected brain regions closer together in a lower-dimensional embedding space, indicating similar functional scores, while regions with limited or no inter-covariance are situated farther apart, indicating dissimilar scores. Previous studies have shown that spatial gradients of RSFC variations obtained from nonlinear dimensionality reduction mirror the presumed cortical hierarchy(Bernhardt et al., 2022; Margulies et al., 2016a; Mesulam, 1998), suggesting that functional gradients may approximate an intrinsic coordinate system of the human cortex. To derive the functional gradient, we retained the top 10% entries for each row and computed a cosine similarity matrix from the average RSFC matrix of our entire sample (combining discovery and replication samples, N=5201), The matrix of similarities was subsequently transformed into a normalized angular matrix to eliminate the presence of negative values (Larivière et al., 2020; Paquola et al., 2019a). The α parameter (set at α = 0.5) modulates the influence of sampling point density on the manifold (α = 0 indicates maximal influence; α = 1 indicates no influence), while the t parameter (set at t = 0) scales eigenvalues of the diffusion operator. These parameters were chosen to preserve the global relationships between data points in the embedded space, as established in previous studies(Hong et al., 2019; Margulies et al., 2016a; Paquola et al., 2019b). Subsequently, the technique of diffusion map embedding was employed to identify the gradient components that account for the variability observed within connectivity pattern of the functional connectome. The primary functional gradient accounted for 27.9% of the resting-state functional connectivity (RSFC) variance, distinguishing primary somatosensory/motor and visual regions from transmodal association cortices, as depicted in Figure S2. This outcome aligns with previous research conducted on RSFC in a cohort of healthy adults, as reported by (Margulies et al., 2016b). Prior studies have indicated that the sensory-to-transmodal gradient emerges as the dominant gradient around the age of 12-13 years, as observed by (Dong et al., 2021).Notably, the second gradient, which contrasts somatosensory/motor and visual areas, explains a similar amount of variance as the first gradient, approximately 26%, as shown in Figure S2. This finding may suggest that the participants in our study have recently transitioned to a more mature functional organization that is more spatially distributed (Dong et al., 2021).

---

**Table S1.** The assessment of pediatric sleep related problems in the present study.

| Item number in the ABCD cohort | Questions | Abbreviations | SRP-domain |
| --- | --- | --- | --- |
| Sleepdisturb1_p | How many hours of sleep does your child get on most nights? | Nightly sleep hours | sleep onset and maintenance |
| Sleepdisturb2_p | How long after going to bed does your child usually fall asleep? | Typical bedtime sleep latency | sleep onset and maintenance |
| Sleepdisturb3_p | The child goes to bed reluctantly. | Hesitates bedtime | sleep onset and maintenance |
| Sleepdisturb4_p | The child has difficulty getting to sleep at night. | Struggles with night sleep | sleep onset and maintenance |
| Sleepdisturb5_p | The child feels anxious or afraid when falling asleep. | Experiences sleep-time anxiety | sleep onset and maintenance |
| Sleepdisturb6_p | The child startles or jerks parts of the body while falling asleep. | Twitches while sleeping | sleep-wake transitions |
| Sleepdisturb7_p | The child shows repetitive actions such as rocking or head banging while fall asleep. | Displays repetitive actions | sleep-wake transitions |
| Sleepdisturb8_p | The child experiences vivid dream-like scenes while falling asleep. | Has vivid pre-sleep dreams | sleep-wake transitions |
| Sleepdisturb9_p | The child sweats excessively while falling asleep. | Excessively sweats when sleeping | sleep hyperhidrosis |
| Sleepdisturb10_p | The child wakes up more than twice per night. | Wakes frequently at night | sleep onset and maintenance |
| Sleepdisturb11_p | After waking up in the night, the child has difficulty to fall asleep again. | Struggles to resume sleep post-waking | sleep onset and maintenance |
| Sleepdisturb12_p | The child has frequent twitching or jerking of legs while asleep or often changes position during the night or kicks the covers off the bed. | Legs twitch while sleeping, changes positions, or kicks off covers | sleep-wake transitions |
| Sleepdisturb13_p | The child has difficulty in breathing during the night. | Experiences nighttime breathing issues | sleep breathing disorders |
| Sleepdisturb14_p | The child gasps for breaths or is unable to breathe during sleep. | Gasps or struggles to breathe while asleep | sleep breathing disorders |
| Sleepdisturb15_p | The child snores. | Child snores | sleep breathing disorders |
| Sleepdisturb16_p | The child sweats excessively during the night. | Sweats excessively at night | sleep hyperhidrosis |
| Sleepdisturb17_p | You have observed the child sleepwalking. | Sleepwalk | problems with arousal |
| Sleepdisturb18_p | You have observed the child talking in their sleep. | Sleep talking | sleep-wake transitions |
| Sleepdisturb19_p | The child grinds their teeth during sleep. | Engages in nocturnal teeth grinding | sleep-wake transitions |
| Sleepdisturb20_p | The child wakes from sleep screaming or confused so you cannot seem to get through to they, but has no memory of these events the next morning. | Wakes screaming or confused, unresponsive, with no memory next morning. | problems with arousal |
| Sleepdisturb21_p | The child has nightmares which they don't remember the next day. | Has forgotten nightmares | problems with arousal |
| Sleepdisturb22_p | The child is unusually difficult to wake up in the morning. | Unusually hard to wake in the morning | excessive somnolence |
| Sleepdisturb23_p | The child awakes in the morning feeling tired | Wakes up tired in the morning | excessive somnolence |
| Sleepdisturb24_p | The child feels unable to move when waking up in the morning. | Experiences morning sleep paralysis | excessive somnolence |
| Sleepdisturb25_p | The child experiences daytime sleepiness. | Has daytime sleepiness | excessive somnolence |
| Sleepdisturb26_p | The child falls asleep suddenly in inappropriate situations. | Experiences sudden inappropriate sleep | excessive somnolence |
| cbcl_q47_p | Nightmares | Nightmares |  |
| cbcl_q54_p | Overtired without good reason | Overtired without good reason |  |
| cbcl_q76_p | Sleeps less than most kids | Sleeps less than most kids |  |
| cbcl_q77_p | Sleeps more than most kids during day and/or night | Sleeps more than most kids during day and/or night |  |
| cbcl_q100_p | Trouble sleeping | Trouble sleeping |  |
| cbcl_q108_p | Wets the bed | Wets the bed |  |

**Note.** ABCD, Adolescent Brain Cognitive Development; cbcl, child behavior checklist; sleepdisturb, sleep disturbance scale for children.

**Table S2.** Pearson’s correlations between the original behavior/imaging loadings and the loadings from control PLS analyses.

|  |  | Site harmonization | PCA explaining 30% variance | PCA explaining 70% variance | Quantile normalization for SRP | T1w manual quality control scores as covariables in structure features | Cortical Volume removal |
| --- | --- | --- | --- | --- | --- | --- | --- |
| PCs | Surface area PCs | 50 | 22 | 104 | 51 | 51 | 51 |
|  | Thickness PCs | 66 | 14 | 151 | 65 | 65 | 65 |
|  | Volume PCs | 55 | 22 | 127 | 58 | 58 | - |
|  | RSFC PCs | 263 | 64 | 738 | 262 | 262 | 262 |
|  | Total PCs | 434 | 122 | 1120 | 436 | 436 | 378 |
| Correlation with loadings from original PLS (LC1) | Behavior loadings | 1 | 1 | 1 | 0.98 | 0.99 | 0.99 |
|  | Area loadings | 0.99 | 0.76 | 0.84 | 0.99 | 0.99 | 0.97 |
|  | Thickness loadings | 0.98 | 0.70 | 0.82 | 0.99 | 0.99 | 0.96 |
|  | Volume loadings | 0.99 | 0.75 | 0.85 | 0.99 | 0.99 | - |
|  | RSFC loadings | 0.99 | 0.76 | 0.86 | 0.99 | 0.99 | 0.98 |
| Correlation with loadings from original PLS (LC3) | Behavior loadings | 0.98 | 0.81 | 0.94 | 0.86 | 0.99 | 0.99 |
|  | Area loadings | 0.98 | 0.52 | 0.83 | 0.83 | 0.99 | 0.97 |
|  | Thickness loadings | 0.98 | 0.38 | 0.77 | 0.83 | 0.99 | 0.96 |
|  | Volume loadings | 0.98 | 0.58 | 0.81 | 0.81 | 0.99 | - |
|  | RSFC loadings | 0.99 | 0.77 | 0.87 | 0.91 | 0.99 | 0.99 |

The number of principal components (PCs) used per imaging modality is specified for each control analysis. Significant correlations are highlighted in bold. Abbreviations: PCA = principal component analysis; PCs = principal components.

**Table S3.** Model fit, intercept and slope for internalizing/externalizing problems.

| Variable | Mode fit |  |  | Growth factors |  |
| --- | --- | --- | --- | --- | --- |
|  | CFI | RMSEA | SRMR | Intercept | Slope |
| IP | 1.000 | 0.011 | 0.004 | 4.956*** | -0.003 |
| EP | 1.000 | 0.000 | 0.000 | 4.087*** | -0.234*** |

**Note.** IP, internalizing problems; EP, externalizing problems; CFI, Comparative Fit Index; RMSEA, Root Mean Square Error of Approximation; SRMR, Standardized Root Mean Square Residual.

**Figure S1.**
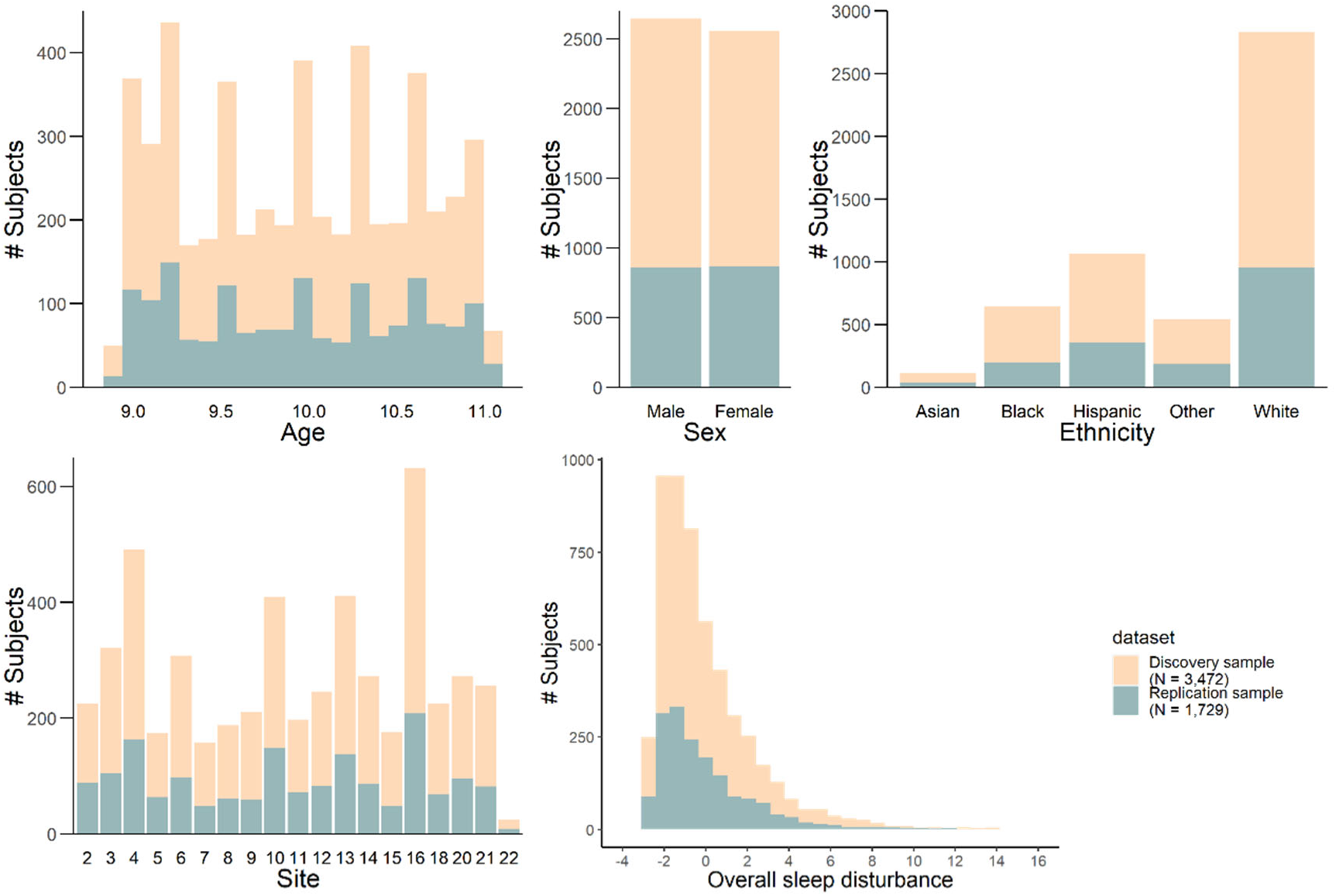
Distribution of social-demographic variables for the discovery and replication datasets.

**Figure S2.**
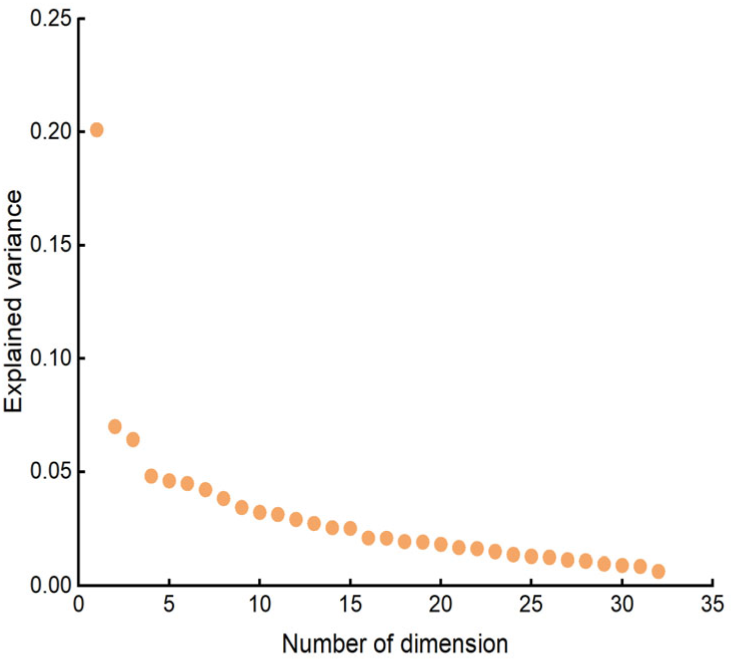
The amount of covariance explained by each latent variable (LV) or latent component (LC). Each orange dot represents a LC.

**Figure S3.**
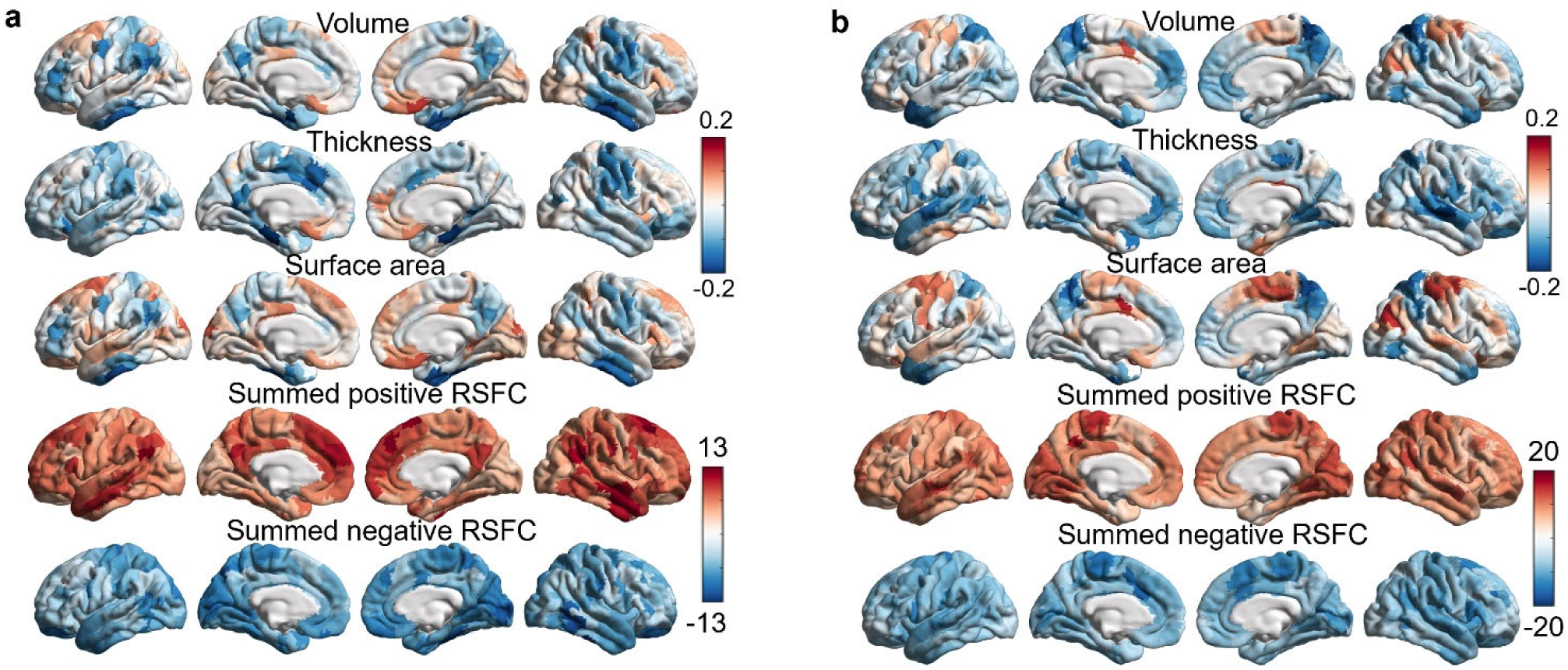
(a) Unthresholded multimodal neural maps of dimension of SPR (LC1). (b) Unthresholded multimodal neural maps of dimension of SPR (LC3)

## Notes

### Competing Interest Statement

The authors have declared no competing interest.

## References

Abi-Dargham, A., Moeller, S. J., Ali, F., DeLorenzo, C., Domschke, K., Horga, G., Jutla, A., Kotov, R., Paulus, M. P., Rubio, J. M., Sanacora, G., Veenstra-VanderWeele, J., & Krystal, J. H. (2023). Candidate biomarkers in psychiatric disorders: state of the field. In World Psychiatry (Vol. 22, Issue 2). 10.1002/wps.21078

Achenbach, T. M., & Edelbrock, C. S. (1983). Manual for the Child Behavior Checklist and revised child behavior profile. University of Vermont Department of Psychiatry.

Akbar, S. A., Mattfeld, A. T., Laird, A. R., & McMakin, D. L. (2022). Sleep to Internalizing Pathway in Young Adolescents (SIPYA): A proposed neurodevelopmental model. Neuroscience and Biobehavioral Reviews, 140(July), 104780. 10.1016/j.neubiorev.2022.104780

Alexander-Bloch, A. F., Shou, H., Liu, S., Satterthwaite, T. D., Glahn, D. C., Shinohara, R. T., Vandekar, S. N., & Raznahan, A. (2018). On testing for spatial correspondence between maps of human brain structure and function. NeuroImage, 178, 540–551. 10.1016/j.neuroimage.2018.05.070

Anastasiades, P. G., de Vivo, L., Bellesi, M., & Jones, M. W. (2022). Adolescent sleep and the foundations of prefrontal cortical development and dysfunction. In Progress in Neurobiology (Vol. 218). 10.1016/j.pneurobio.2022.102338

Bonuck, K., Freeman, K., Chervin, R. D., & Xu, L. (2012). Sleep-disordered breathing in a population-based cohort: Behavioral outcomes at 4 and 7 years. Pediatrics, 129(4). 10.1542/peds.2011-1402

Bruni, O., Ottaviano, S., Guidetti, V., Romoli, M., Innocenzi, M., Cortesi, F., & Giannotti, F. (1996). The Sleep Disturbance Scale for Children (SDSC) construction and validation of an instrument to evaluate sleep disturbances in childhood and adolescence. Journal of Sleep Research, 5(4), 251–261. 10.1111/j.1365-2869.1996.00251.x

Burt, J. B., Helmer, M., Shinn, M., Anticevic, A., & Murray, J. D. (2020). Generative modeling of brain maps with spatial autocorrelation. NeuroImage, 220(June), 117038. 10.1016/j.neuroimage.2020.117038

Casey, B. J., Cannonier, T., Conley, M. I., Cohen, A. O., Barch, D. M., Heitzeg, M. M., Soules, M. E., Teslovich, T., Dellarco, D. V., Garavan, H., Orr, C. A., Wager, T. D., Banich, M. T., Speer, N. K., Sutherland, M. T., Riedel, M. C., Dick, A. S., Bjork, J. M., Thomas, K. M., … Dale, A. M. (2018). The Adolescent Brain Cognitive Development (ABCD) study: Imaging acquisition across 21 sites. In Developmental Cognitive Neuroscience (Vol. 32). 10.1016/j.dcn.2018.03.001

Chen, M., Li, Y., Chen, J., Gao, L., Sun, J., Gu, Z., Wu, T., & Chan, P. (2022). Structural and functional brain alterations in patients with idiopathic rapid eye movement sleep behavior disorder. Journal of Neuroradiology, 49(1). 10.1016/j.neurad.2020.04.007

Cheng, W., Rolls, E., Gong, W., Du, J., Zhang, J., Zhang, X. Y., Li, F., & Feng, J. (2021). Sleep duration, brain structure, and psychiatric and cognitive problems in children. Molecular Psychiatry, 26(8), 3992–4003. 10.1038/s41380-020-0663-2

Chiang, J. J., Lam, P. H., Chen, E., & Miller, G. E. (2022). Psychological Stress During Childhood and Adolescence and Its Association With Inflammation Across the Lifespan: A Critical Review and Meta-Analysis. Psychological Bulletin, 148(1–2). 10.1037/bul0000351

Cooper, R., Di Biase, M. A., Bei, B., Quach, J., & Cropley, V. (2023). Associations of Changes in Sleep and Emotional and Behavioral Problems from Late Childhood to Early Adolescence. JAMA Psychiatry, 80(6). 10.1001/jamapsychiatry.2023.0379

Corkum, P. V, Allen, S. L., Howlett, M. D., & Aim, J. (2016). ABCs of SLEEPING : A review of the evidence behind pediatric sleep practice recommendations. Sleep Medicine Reviews, 29, 1–14. 10.1016/j.smrv.2015.08.006

Crowley, S. J., Wolfson, A. R., Tarokh, L., & Carskadon, M. A. (2018). An update on adolescent sleep: New evidence informing the perfect storm model. Journal of Adolescence, 67, 55–65.

Dennis, E., Manza, P., & Volkow, N. D. (2022). Socioeconomic status, BMI, and brain development in children. Translational Psychiatry, 12(1). 10.1038/s41398-022-01779-3

Dong, H., Margulies, D. S., Zuo, X., & Holmes, A. J. (2021). Shifting gradients of macroscale cortical organization mark the transition from childhood to adolescence. Proceedings of National Academy of Sciences, USA, 118(28), e2024448118. 10.1073/pnas.2024448118

Dong, H.-M., Marguliesc, D. S., Zuo, X.-N., & Holmes, A. J. (2021). Shifting gradients of macroscale cortical organization mark the transition from childhood to adolescence. Proceedings of the National Academy of Sciences of the United States of America, 118(28).

Dukart, J., Holiga, S., Rullmann, M., Hawkins, P. C. T., Mehta, M. A., Hesse, S., Barthel, H., Sabri, O., Jech, R., & Eickhoff, S. B. (2021). JuSpace : A tool for spatial correlation analyses of magnetic resonance imaging data with nuclear imaging derived neurotransmitter maps. Human Brain Mapping, 42(3), 555–566. 10.1002/hbm.25244

Duncan, T. E., Duncan, S. C., & Strycker, L. A. (2013). An introduction to latent variable growth curve modeling: Concepts, issues, and applications. In An Introduction to Latent Variable Growth Curve Modeling: Concepts, Issues, and Application, Second Edition. 10.4324/9780203879962

Dutil, C., Walsh, J. J., Featherstone, R. B., Gunnell, K. E., Tremblay, M. S., Gruber, R., Weiss, S. K., Cote, K. A., Sampson, M., & Chaput, J. P. (2018). Influence of sleep on developing brain functions and structures in children and adolescents: A systematic review. In Sleep Medicine Reviews (Vol. 42). 10.1016/j.smrv.2018.08.003

Elliott, M. L., Romer, A., Knodt, A. R., & Hariri, A. R. (2018). A Connectome-wide Functional Signature of Transdiagnostic Risk for Mental Illness. Biological Psychiatry, 84(6), 452–459. 10.1016/j.biopsych.2018.03.012

Fischl, B., Salat, D. H., Busa, E., Albert, M., Dieterich, M., Haselgrove, C., Van Der Kouwe, A., Killiany, R., Kennedy, D., Klaveness, S., Montillo, A., Makris, N., Rosen, B., & Dale, A. M. (2002). Whole brain segmentation: Automated labeling of neuroanatomical structures in the human brain. Neuron, 33(3), 341–355. 10.1016/S0896-6273(02)00569-X

Fischl, B., Sereno, M. I., & Dale, A. M. (1999). Cortical surface-based analysis: II. Inflation, flattening, and a surface-based coordinate system. NeuroImage, 9(2). 10.1006/nimg.1998.0396

Fortin, J. P., Cullen, N., Sheline, Y. I., Taylor, W. D., Aselcioglu, I., Cook, P. A., Adams, P., Cooper, C., Fava, M., McGrath, P. J., McInnis, M., Phillips, M. L., Trivedi, M. H., Weissman, M. M., & Shinohara, R. T. (2018). Harmonization of cortical thickness measurements across scanners and sites. NeuroImage, 167. 10.1016/j.neuroimage.2017.11.024

Freeman, D., Sheaves, B., Waite, F., Harvey, A. G., & Harrison, P. J. (2020). Sleep disturbance and psychiatric disorders. In The Lancet Psychiatry (Vol. 7, Issue 7). 10.1016/S2215-0366(20)30136-X

Friedman, N. P., Corley, R. P., Hewitt, J. K., & Wright, K. P. (2009). Individual differences in childhood sleep problems predict later cognitive executive control. Sleep, 32(3), 323–333. 10.1093/sleep/32.3.323

Friel, P., Duran, A. T., Shechter, A., & Diaz, K. M. (2020). U.S. Children Meeting Physical Activity, Screen Time, and Sleep Guidelines. American Journal of Preventive Medicine, 59(4), 513–521. 10.1016/j.amepre.2020.05.007

Galván, A. (2020). The Need for Sleep in the Adolescent Brain. In Trends in Cognitive Sciences (Vol. 24, Issue 1). 10.1016/j.tics.2019.11.002

Garavan, H., Bartsch, H., Conway, K., Decastro, A., Goldstein, R. Z., Heeringa, S., Jernigan, T., Potter, A., Thompson, W., & Zahs, D. (2018). Recruiting the ABCD sample: Design considerations and procedures. Developmental Cognitive Neuroscience, 32. 10.1016/j.dcn.2018.04.004

Giddens, N. T., Juneau, P., Manza, P., Wiers, C. E., & Volkow, N. D. (2022). Disparities in sleep duration among American children: effects of race and ethnicity, income, age, and sex. Proceedings of the National Academy of Sciences of the United States of America, 119(30). 10.1073/pnas.2120009119

Glasser, M. F., Coalson, T. S., Robinson, E. C., Hacker, C. D., Harwell, J., Yacoub, E., Ugurbil, K., Andersson, J., Beckmann, C. F., Jenkinson, M., Smith, S. M., & Van Essen, D. C. (2016). A multi-modal parcellation of human cerebral cortex. Nature, 536(7615), 171–178. 10.1038/nature18933

Gregory, A. M., & Sadeh, A. (2016). Annual Research Review: Sleep problems in childhood psychiatric disorders - A review of the latest science. In Journal of Child Psychology and Psychiatry and Allied Disciplines (Vol. 57, Issue 3). 10.1111/jcpp.12469

Hagler, D. J., Hatton, S. N., Cornejo, M. D., Makowski, C., Fair, D. A., Dick, A. S., Sutherland, M. T., Casey, B. J., Barch, D. M., Harms, M. P., Watts, R., Bjork, J. M., Garavan, H. P., Hilmer, L., Pung, C. J., Sicat, C. S., Kuperman, J., Bartsch, H., Xue, F., … Dale, A. M. (2019). Image processing and analysis methods for the Adolescent Brain Cognitive Development Study. NeuroImage, 202, 116091. 10.1016/j.neuroimage.2019.116091

Haritos, R., Küppers, V., Samea, F., Riemann, D., Jessen, F., Eickhoff, S. B., Dafsari, F. S., & Tahmasian, M. (2025). The effect of psychotherapy on the multivariate association between insomnia and depressive symptoms in late-life depression. 10.1101/2025.03.28.25324836

Hayes, A. F. (2009). Beyond Baron and Kenny: Statistical Mediation Analysis in the New Millennium. Communication Monographs, 76(4), 408–420.

Hill, J., Inder, T., Neil, J., Dierker, D., Harwell, J., & Van Essen, D. (2010). Similar patterns of cortical expansion during human development and evolution. Proceedings of the National Academy of Sciences of the United States of America, 107(29), 13135–13140. 10.1073/pnas.1001229107

Hill, Kashyap, P., Raffanello, E., Wang, Y., Moffitt, T. E., Caspi, A., Engelhard, M., & Posner, J. (2025). Prediction of mental health risk in adolescents. Nature Medicine, 1–7.

Hubbard, N. A., Siless, V., Frosch, I. R., Goncalves, M., Lo, N., Wang, J., Bauer, C. C. C., Conroy, K., Cosby, E., Hay, A., Jones, R., Pinaire, M., Vaz De Souza, F., Vergara, G., Ghosh, S., Henin, A., Hirshfeld-Becker, D. R., Hofmann, S. G., Rosso, I. M., … Whitfield-Gabrieli, S. (2020). Brain function and clinical characterization in the Boston adolescent neuroimaging of depression and anxiety study. NeuroImage: Clinical, 27(March). 10.1016/j.nicl.2020.102240

Huntenburg, J. M., Bazin, P. L., & Margulies, D. S. (2018). Large-Scale Gradients in Human Cortical Organization. Trends in Cognitive Sciences, 22(1), 21–31. 10.1016/j.tics.2017.11.002

Hysing, M., Harvey, A. G., Bøe, T., Heradstveit, O., Vedaa, Ø., & Sivertsen, B. (2020). Trajectories of sleep problems from adolescence to adulthood. Linking two population-based studies from Norway. Sleep Medicine, 75, 411–417. 10.1016/j.sleep.2020.08.035

Isaiah, A., Ernst, T., Cloak, C. C., Clark, D. B., & Chang, L. (2021). Associations between frontal lobe structure, parent-reported obstructive sleep disordered breathing and childhood behavior in the ABCD dataset. Nature Communications, 12(2205), 1–10. 10.1038/s41467-021-22534-0

Laurent, J. S., Watts, R., Adise, S., Allgaier, N., Chaarani, B., Garavan, H., Potter, A., & Mackey, S. (2020). Associations among Body Mass Index, Cortical Thickness, and Executive Function in Children. JAMA Pediatrics, 174(2). 10.1001/jamapediatrics.2019.4708

Leong, R. L. F., & Chee, M. W. L. (2023). Understanding the Need for Sleep to Improve Cognition. In Annual Review of Psychology (Vol. 74). 10.1146/annurev-psych-032620-034127

Lo, J. C., Ong, J. L., Leong, R. L. F., Gooley, J. J., & Chee, M. W. L. (2016). Cognitive performance, sleepiness, and mood in partially sleep deprived adolescents: The need for sleep Study. Sleep, 39(3). 10.5665/sleep.5552

Luo, A. C., Sydnor, V. J., Pines, A., Larsen, B., Alexander-Bloch, A. F., Cieslak, M., Covitz, S., Chen, A. A., Esper, N. B., Feczko, E., Franco, A. R., Gur, R. E., Gur, R. C., Houghton, A., Hu, F., Keller, A. S., Kiar, G., Mehta, K., Salum, G. A., … Satterthwaite, T. D. (2024). Functional connectivity development along the sensorimotor-association axis enhances the cortical hierarchy. Nature Communications, 15(1). 10.1038/s41467-024-47748-w

Markello, R. D., Hansen, J. Y., Liu, Z. Q., Bazinet, V., Shafiei, G., Suárez, L. E., Blostein, N., Seidlitz, J., Baillet, S., Satterthwaite, T. D., Chakravarty, M. M., Raznahan, A., & Misic, B. (2022). neuromaps: structural and functional interpretation of brain maps. Nature Methods, 19(11). 10.1038/s41592-022-01625-w

Markello, R. D., & Misic, B. (2021). Comparing spatial null models for brain maps. NeuroImage, 236(August 2020), 118052. 10.1016/j.neuroimage.2021.118052

Mason, G. M., Lokhandwala, S., Riggins, T., & Spencer, R. M. C. (2021). Sleep and human cognitive development. In Sleep Medicine Reviews (Vol. 57). 10.1016/j.smrv.2021.101472

McCurry, K. L., Toda-Thorne, K., Taxali, A., Angstadt, M., Hardi, F. A., Heitzeg, M. M., & Sripada, C. (2024). Data-driven, generalizable prediction of adolescent sleep disturbances in the multisite Adolescent Brain Cognitive Development Study. SLEEP. 10.1093/sleep/zsae048

McIntosh, A. R., & Lobaugh, N. J. (2004). Partial least squares analysis of neuroimaging data: Applications and advances. NeuroImage, 23, S250–S263. 10.1016/j.neuroimage.2004.07.020

McIntosh, A. R., & Mišić, B. (2013). Multivariate statistical analyses for neuroimaging data. Annual Review of Psychology, 64, 499–525. 10.1146/annurev-psych-113011-143804

Melaku, Y. A., Appleton, S., Reynolds, A. C., Sweetman, A. M., Stevens, D. J., Lack, L., & Adams, R. (2019). Association between Childhood Behavioral Problems and Insomnia Symptoms in Adulthood. JAMA Network Open, 2(9), 1–17. 10.1001/jamanetworkopen.2019.10861

Meltzer, L. J., Williamson, A. A., & Mindell, J. A. (2021). Pediatric sleep health: It matters, and so does how we define it. Sleep Medicine Reviews, 57, 1–30. 10.1016/j.smrv.2021.101425

Meng, S. Q., Cheng, J. L., Li, Y. Y., Yang, X. Q., Zheng, J. W., Chang, X. W., Shi, Y., Chen, Y., Lu, L., Sun, Y., Bao, Y. P., & Shi, J. (2022). Global prevalence of digital addiction in general population: A systematic review and meta-analysis. In Clinical Psychology Review (Vol. 92). 10.1016/j.cpr.2022.102128

Menon, V. (2023). 20 years of the default mode network: A review and synthesis. In Neuron (Vol. 111, Issue 16). 10.1016/j.neuron.2023.04.023

Morales-Muñoz, I., & Gregory, A. M. (2023). Sleep and Mental Health Problems in Children and Adolescents. In Sleep Medicine Clinics (Vol. 18, Issue 2). 10.1016/j.jsmc.2023.01.006

Morrissey, B., Taveras, E., Allender, S., & Strugnell, C. (2020). Sleep and obesity among children: A systematic review of multiple sleep dimensions. In Pediatric Obesity (Vol. 15, Issue 4). 10.1111/ijpo.12619

Palmer, C. A., Bower, J. L., Cho, K. W., Clementi, M. A., Lau, S., Oosterhoff, B., & Alfano, C. A. (2023). Sleep Loss and Emotion: A Systematic Review and Meta-Analysis of Over 50 Years of Experimental Research. Psychological Bulletin, 150(4). 10.1037/bul0000410

Porkka-Heiskanen, T., Zitting, K. M., & Wigren, H. K. (2013). Sleep, its regulation and possible mechanisms of sleep disturbances. In Acta Physiologica (Vol. 208, Issue 4). 10.1111/apha.12134

Reardon, P. K., Seidlitz, J., Vandekar, S., Liu, S., Patel, R., Tae, M., Park, M., Alexander-Bloch, A., Clasen, L. S., Blumenthal, J. D., Lalonde, F. M., Giedd, J. N., Gur, R. C., Gur, R. E., Lerch, J. P., Chakravarty, M. M., Satterthwaite, T. D., Shinohara, R. T., & Raznahan, A. (2018). Normative brain size variation and brain shape diversity in humans. Science, 360(6394), 1222–1227. https://www.science.org

Reynolds, A. M., Spaeth, A. M., Hale, L., Williamson, A. A., LeBourgeois, M. K., Wong, S. D., Hartstein, L. E., Levenson, J. C., Kwon, M., Hart, C. N., Greer, A., Richardson, C. E., Gradisar, M., Clementi, M. A., Simon, S. L., Reuter-Yuill, L. M., Picchietti, D. L., Wild, S., Tarokh, L., … Carskadon, M. A. (2023). Pediatric sleep: current knowledge, gaps, and opportunities for the future. Sleep, 46(7), 1–21. 10.1093/sleep/zsad060

Ribeiro, M., Yordanova, Y. N., Noblet, V., Herbet, G., & Ricard, D. (2024). White matter tracts and executive functions: A review of causal and correlation evidence. In Brain (Vol. 147, Issue 2). 10.1093/brain/awad308

Ricci, A., & Fernandez-Mendoza, J. (2024). Evidence of how the maturing sleeping brain contributes to the sleepy brain of adolescents. In Sleep (Vol. 47, Issue 1). 10.1093/sleep/zsad283

Riemann, D., Spiegelhalder, K., Feige, B., Voderholzer, U., Berger, M., Perlis, M., & Nissen, C. (2010). The hyperarousal model of insomnia: a review of the concept and its evidence. Sleep Medicine Reviews, 14(1), 19–31.

Royer, J., Kebets, V., Piguet, C., Chen, J., Ooi, L. Q. R., Kirschner, M., Siffredi, V., Misic, B., Thomas Yeo, B. T., & Bernhardt, B. C. (2024). MULTIMODAL NEURAL CORRELATES OF CHILDHOOD PSYCHOPATHOLOGY. In elife. 10.1101/2023.03.02.530821

Royer, J., Kebets, V., Piguet, C., Chen, J., Rong, L. Q., Kirschner, M., Siffredi, V., Misic, B., Thomas, B. T., Bernhardt, B. C., & Bernhardt, B. (2024). Multimodal neural correlates of childhood psychopathology. ELife, 13(e87992).

Schaefer, A., Kong, R., Gordon, E. M., Laumann, T. O., Zuo, X.-N., Holmes, A. J., Eickhoff, S. B., & Yeo, B. T. T. (2018). Local-Global Parcellation of the Human Cerebral Cortex from Intrinsic Functional Connectivity MRI. Cerebral Cortex, 28(9), 3095–3114. 10.1093/cercor/bhx179

Scott, J., Kallestad, H., Vedaa, O., Sivertsen, B., & Etain, B. (2021). Sleep disturbances and first onset of major mental disorders in adolescence and early adulthood: A systematic review and meta-analysis. In Sleep Medicine Reviews (Vol. 57). 10.1016/j.smrv.2021.101429

Shen, C., Luo, Q., Chamberlain, S. R., Morgan, S., Romero-Garcia, R., Du, J., Zhao, X., Touchette, É., Montplaisir, J., Vitaro, F., Boivin, M., Tremblay, R. E., Zhao, X. M., Robaey, P., Feng, J., & Sahakian, B. J. (2020). What Is the Link Between Attention-Deficit/Hyperactivity Disorder and Sleep Disturbance? A Multimodal Examination of Longitudinal Relationships and Brain Structure Using Large-Scale Population-Based Cohorts. Biological Psychiatry, 88(6). 10.1016/j.biopsych.2020.03.010

Simon, K. C., Nadel, L., & Payne, J. D. (2022). The functions of sleep: A cognitive neuroscience perspective. Proceedings of the National Academy of Sciences of the United States of America, 119(44). 10.1073/pnas.2201795119

Somerville, L. H., Bookheimer, S. Y., Buckner, R. L., Burgess, G. C., Curtiss, S. W., Dapretto, M., Elam, J. S., Gaffrey, M. S., Harms, M. P., Hodge, C., Kandala, S., Kastman, E. K., Nichols, T. E., Schlaggar, B. L., Smith, S. M., Thomas, K. M., Yacoub, E., Van Essen, D. C., & Barch, D. M. (2018). The Lifespan Human Connectome Project in Development: A large-scale study of brain connectivity development in 5–21 year olds. NeuroImage, 183, 456–468. 10.1016/j.neuroimage.2018.08.050

Sydnor, V. J., Larsen, B., Bassett, D. S., Alexander-Bloch, A., Fair, D. A., Liston, C., Mackey, A. P., Milham, M. P., Pines, A., Roalf, D. R., Seidlitz, J., Xu, T., Raznahan, A., & Satterthwaite, T. D. (2021). Neurodevelopment of the association cortices: Patterns, mechanisms, and implications for psychopathology. In Neuron (Vol. 109, Issue 18). 10.1016/j.neuron.2021.06.016

Telzer, E. H., Fuligni, A. J., Lieberman, M. D., & Galván, A. (2013). The effects of poor quality sleep on brain function and risk taking in adolescence. NeuroImage, 71. 10.1016/j.neuroimage.2013.01.025

Thomas, A. G., Monahan, K. C., Lukowski, A. F., & Cauffman, E. (2015). Sleep Problems Across Development: A Pathway to Adolescent Risk Taking Through Working Memory. Journal of Youth and Adolescence, 44(2), 447–464. 10.1007/s10964-014-0179-7

Turan, O., Garner, J., Isaiah, A., Palatino, M., Ernst, T., Wang, Z., & Chang, L. (2025). Fitbit-Measured Sleep Duration in Young Adolescents is Associated with Functional Connectivity in Attentional, Executive Control, Memory, and Sensory Networks. Sleep, zsaf088.

Váša, F., Seidlitz, J., Romero-Garcia, R., Whitaker, K. J., Rosenthal, G., Vértes, P. E., Shinn, M., Alexander-Bloch, A., Fonagy, P., Dolan, R. J., Jones, P. B., Goodyer, I. M., Sporns, O., & Bullmore, E. T. (2018). Adolescent tuning of association cortex in human structural brain networks. Cerebral Cortex, 28(1). 10.1093/cercor/bhx249

Volkow, N. D., Koob, G. F., Croyle, R. T., Bianchi, D. W., Gordon, J. A., Koroshetz, W. J., Pérez-stable, E. J., Riley, W. T., Bloch, M. H., Conway, K., Deeds, B. G., Dowling, G. J., Grant, S., Howlett, K. D., Matochik, J. A., Morgan, G. D., Murray, M. M., Noronha, A., Spong, C. Y., … Weiss, S. R. B. (2018). The conception of the ABCD study : From substance use to a broad NIH collaboration. Developmental Cognitive Neuroscience, 32, 4–7. 10.1016/j.dcn.2017.10.002

Willett, J. B., & Sayer, A. G. (1994). Using Covariance Structure Analysis to Detect Correlates and Predictors of Individual Change Over Time. Psychological Bulletin, 116(2). 10.1037//0033-2909.116.2.363

Xia, C. H., Ma, Z., Ciric, R., Gu, S., Betzel, R. F., Kaczkurkin, A. N., Calkins, M. E., Cook, P. A., García de la Garza, A., Vandekar, S. N., Cui, Z., Moore, T. M., Roalf, D. R., Ruparel, K., Wolf, D. H., Davatzikos, C., Gur, R. C., Gur, R. E., Shinohara, R. T., … Satterthwaite, T. D. (2018). Linked dimensions of psychopathology and connectivity in functional brain networks. Nature Communications, 9(1), 1–14. 10.1038/s41467-018-05317-y

Yeo, B. T. T., Krienen, F. M., Sepulcre, J., Sabuncu, M. R., Lashkari, D., Hollinshead, M., Roffman, J. L., Smoller, J. W., Zöllei, L., Polimeni, J. R., Fischl, B., Liu, H., & Buckner, R. L. (2011). The organization of the human cerebral cortex estimated by intrinsic functional connectivity. Journal of Neurophysiology, 106, 1125–1165. 10.1152/jn.00338.2011.

Yeshurun, Y., Nguyen, M., & Hasson, U. (2021). The default mode network: where the idiosyncratic self meets the shared social world. Nature Reviews Neuroscience, 22(3). 10.1038/s41583-020-00420-w

Zhao, Y., Yang, L., Sahakian, B. J., Langley, C., Zhang, W., Kuo, K., Li, Z., Gan, Y., Li, Y., Zhao, Y., Yu, J., Feng, J., & Cheng, W. (2023). The brain structure, immunometabolic and genetic mechanisms underlying the association between lifestyle and depression. Nature Mental Health, 1(10). 10.1038/s44220-023-00120-1

Zhi, D., Jiang, R., Pearlson, G., Fu, Z., Qi, S., Yan, W., Feng, A., Xu, M., Calhoun, V., & Sui, J. (2024). Triple Interactions Between the Environment, Brain, and Behavior in Children: An ABCD Study. Biological Psychiatry, 95(9). 10.1016/j.biopsych.2023.12.019

Zöller, D., Schaer, M., Scariati, E., Padula, M. C., Eliez, S., & Van De Ville, D. (2017). Disentangling resting-state BOLD variability and PCC functional connectivity in 22q11.2 deletion syndrome. NeuroImage, 149, 85–97. 10.1016/j.neuroimage.2017.01.064

## References in this supplement materials

Bernhardt, B. C., Smallwood, J., Keilholz, S., & Margulies, D. S. (2022). Gradients in brain organization. In NeuroImage (Vol. 251). Academic Press Inc. 10.1016/j.neuroimage.2022.118987

Coifman, R. R., Lafon, S., Lee, A. B., Maggioni, M., Nadler, B., Warner, F., & Zucker, S. W. (2005). Geometric diffusions as a tool for harmonic analysis and structure definition of data: Diffusion maps. Proceedings of the National Academy of Sciences of the United States of America, 102(21), 7426–7431. 10.1073/pnas.0500334102

Fischl, B., Sereno, M. I., Tootell, R. B. H., & Dale, A. M. (1999). High-resolution intersubject averaging and a coordinate system for the cortical surface. Human Brain Mapping, 8(4). 10.1002/(SICI)1097-0193(1999)8:4<272::AID-HBM10>3.0.CO;2-4

Greve, D. N., & Fischl, B. (2009). Accurate and robust brain image alignment using boundary-based registration. NeuroImage, 48(1). 10.1016/j.neuroimage.2009.06.060

Hagler, D. J., Hatton, S. N., Cornejo, M. D., Makowski, C., Fair, D. A., Dick, A. S., Sutherland, M. T., Casey, B. J., Barch, D. M., Harms, M. P., Watts, R., Bjork, J. M., Garavan, H. P., Hilmer, L., Pung, C. J., Sicat, C. S., Kuperman, J., Bartsch, H., Xue, F., … Dale, A. M. (2019). Image processing and analysis methods for the Adolescent Brain Cognitive Development Study. NeuroImage, 202. 10.1016/j.neuroimage.2019.116091

Hong, S. J., de Wael, R. V., Bethlehem, R. A. I., Lariviere, S., Paquola, C., Valk, S. L., Milham, M. P., Di Martino, A., Margulies, D. S., Smallwood, J., & Bernhardt, B. C. (2019). Atypical functional connectome hierarchy in autism. Nature Communications, 10(1). 10.1038/s41467-019-08944-1

Jenkinson, M., Bannister, P., Brady, M., & Smith, S. (2002). Improved optimization for the robust and accurate linear registration and motion correction of brain images. NeuroImage, 17(2). 10.1016/S1053-8119(02)91132-8

Kong, R., Li, J., Orban, C., Sabuncu, M. R., Liu, H., Schaefer, A., Sun, N., Zuo, X. N., Holmes, A. J., Eickhoff, S. B., & Yeo, B. T. T. (2019). Spatial Topography of Individual-Specific Cortical Networks Predicts Human Cognition, Personality, and Emotion. Cerebral Cortex, 29(6). 10.1093/cercor/bhy123

Larivière, S., Wael, R. V. de, Hong, S.-J., Paquola, C., Tavakol, S., Lowe, A. J., Schrader, D. V, & Bernhardt, B. C. (2020). Multiscale Structure–Function Gradients in the Neonatal Connectome. Cerebral Cortex, 30(1), 47–58.

Margulies, D. S., Ghosh, S. S., Goulas, A., Falkiewicz, M., Huntenburg, J. M., Langs, G., Bezgin, G., Eickhoff, S. B., Castellanos, F. X., Petrides, M., Jefferies, E., & Smallwood, J. (2016a). Situating the default-mode network along a principal gradient of macroscale cortical organization. Proceedings of the National Academy of Sciences of the United States of America, 113(44), 12574–12579. 10.1073/pnas.1608282113

Margulies, D. S., Ghosh, S. S., Goulas, A., Falkiewicz, M., Huntenburg, J. M., Langs, G., Bezgin, G., Eickhoff, S. B., Castellanos, F. X., Petrides, M., Jefferies, E., & Smallwood, J. (2016b). Situating the default-mode network along a principal gradient of macroscale cortical organization. Proceedings of the National Academy of Sciences, 113(44), 12574–12579. 10.1073/pnas.1608282113

Mesulam, M.-M. (1998). From sensation to cognition. Brain, 121, 1013–1052.

Paquola, C., Vos De Wael,R., Wagstyl, K., Bethlehem, R. A. I., Hong, S. J., Seidlitz, J., Bullmore, E. T., Evans, A. C., Misic, B., Margulies, D. S., Smallwood, J., & Bernhardt, B. C. (2019a). Microstructural and functional gradients are increasingly dissociated in transmodal cortices. PLoS Biology, 17(5), 1–28. 10.1371/journal.pbio.3000284

Paquola, C., Vos De Wael, R., Wagstyl, K., Bethlehem, R. A. I., Hong, S. J., Seidlitz, J., Bullmore, E. T., Evans, A. C., Misic, B., Margulies, D. S., Smallwood, J., & Bernhardt, B. C. (2019b). Microstructural and functional gradients are increasingly dissociated in transmodal cortices. PLoS Biology, 17(5). 10.1371/journal.pbio.3000284

Power, J. D., Barnes, K. A., Snyder, A. Z., Schlaggar, B. L., & Petersen, S. E. (2012). Spurious but systematic correlations in functional connectivity MRI networks arise from subject motion. NeuroImage, 59(3), 2142–2154. 10.1016/j.neuroimage.2011.10.018

Power, J. D., Mitra, A., Laumann, T. O., Snyder, A. Z., Schlaggar, B. L., & Petersen, S. E. (2014). Methods to detect, characterize, and remove motion artifact in resting state fMRI. NeuroImage, 84. 10.1016/j.neuroimage.2013.08.048

